# Accuracy and precision of visual acuity tests: A systematic review and meta-analysis across testing strategies

**DOI:** 10.64898/2026.11.04.26366580

**Authors:** Clare Chen, Andrew W. Browne, Jan Skerswetat

## Abstract

**Purpose:** Visual acuity (VA) testing has expanded from examiner-guided to examinee self-guided assessments, yet measurement variability may limit clinical utility. This meta-analysis investigated VA-tests’ intertest and test-retest accuracy and precision across different testing strategies.

**Methods:** Following PRISMA guidelines, we searched PubMed, Google Scholar, and Scopus for VA validation studies published between 1 January 1983 to 02 December 2024. Eligible studies reported intertest or test-retest Bland-Altman mean bias and limits of agreement (LoA); objective VA assessments and correlation coefficient only studies were excluded. Testing strategies were classified by test site, optotype sampling methods, response input methods, and level of supervision. Accuracy outcomes were reported as mean biases. Precision outcomes were reported as intertest LoA half-width and test-retest coefficient of repeatability (CoR) for test-retest. Multilevel random-effects models accounted for multiple estimates per study. Risk of bias was assessed using QUADAS-2.

**Results:** Across 82 included studies (10,873 participants), pooled intertest and test-retest mean biases were both 0.01 logMAR. Intertest LoA half-width was 0.19 logMAR and test-retest CoR was 0.14 logMAR. Variability showed marginal difference by optotype sampling (adaptive: LoA half-width 0.20 logMAR, CoR 0.15 logMAR; fixed: 0.18, 0.13) or response input method (examiner: 0.19, 0.13; self-run: 0.19, 0.18). Only few studies reported results for test site and level of supervision, hence only intertest was compared (test site, clinic: LoA half-width 0.19 logMAR vs remote 0.27 logMAR and level of supervision, supervised: 0.19 logMAR vs unsupervised 0.26 logMAR). QUADAS-2 identified patient selection as the domain of greatest bias concern.

**Conclusion:** Across testing strategies, VA assessments showed minimal systematic bias but substantial variability. Lack of precision is more notable in remote and unsupervised testing while no difference within optotype sampling and response-input methods. Future development should prioritize on improving precision of visual acuity tests with finer adaptive optotype resolution and remote/unsupervised testings through repeatability studies.

## Introduction

Globally, 2.2 billion people have visual impairment, with a substantial impact on development and quality of life (1). Visual acuity (VA) is the primary measure of vision in ophthalmology and optometry (2,3), underpinning disease monitoring, treatment evaluation, and clinical trial endpoints(4). The Earyl Treatment Diabetic Retinopathy Study (ETDRS) chart is widely used as a reference standard in rigorous clinical research(5). However, routine VA testing remains heterogenous, ranging from conventional charts, such as Snellen and LEA charts, to examiner-guided and self-administered adaptive digital tests. Software and mobile platforms(6–10) have further extended testing beyond traditional clinical settings, including patient’s homes and emergency departments(11–13). This diversity in chart design, administration, testing environment, and patient characteristics raises a fundamental question: do these approaches yield VA measurements that are reproducible and comparable?

### Methodologies in Assessing VA

VA is the primary outcome measure in optometric and ophthalmic clinical care and science. Various ways to measure it have been established since its earliest application(14). Conventionally, VA is measured at a dedicated eyecare facility using a wall-mounted or retro-illuminated chart 6 meters from the observer to reduce the influence of near-accommodation and pupil constriction reflex. The administrator then proctors the exam and scores the performance as smallest legible line as logMAR, decimal, or Snellen notation(5). Modifications in psychophysical procedures, optotypes, viewing distances, and administration strategies permit VA tests to be performed within a clinical environment, or remotely, without administrator’s proctoring and scoring(8,10) This expansion in *where* VA is measured has been matched by variation in *how,* namely the psychophysical procedure and scoring convention. Fixed presentation shows every optotype on every line and has fixed size change steps between line of optotypes, e.g., ETDRS 0.10 logMAR. Adaptive procedures instead select each subsequent optotype based on prior response using staircase procedures, Bayesian algorithms, or adaptive charts(10,15). These tests enable estimations of threshold in a customized way and may have finer test resolution (i.e. smaller step sizes between rendered stimuli(10,16)) compared to ETDRS’s 0.10 logMAR step size. Moreover, these tests may provide additional information about the psychophysical performance compared to chart-based tasks (e.g., slope of psychometric function)(17).

### Sources of measurement noise

#### Test Site

Routinely VA is measured in a dedicated eye-care facility where the testing environment is usually held constant. Testing distance is determined by the room set-up (e.g., fixed distance between the examination chair and eye chart) with ambient illumination and chart luminance maintained within recommend ranges per ISO 8596 guidelines(18) or ETDRS protocol(5,19). Additionally, glare and visual distractions were usually minimized. Since these conditions are unlikely to change within the same clinic, patients who consistently return to the same clinic for follow-up visits are less subjected to variability introduced by the different environments. However, if test were performed on different clinical sites, variation in any of these settings could contribute to measurement variability.

However, not all VA assessments were performed in an eye-clinic. For example, patients in the emergency department with visual concerns will need to be evaluated for visual acuity. In the United States, passing a driver’s license test also require checking of VA at the business office, instead of a dedicate eye clinic. During COVID-19, a surge in development of remote VA testing further allows at-home testing(20,21). Unlike eye clinics, these testing environments are less consistent. Viewing distance can be established with facial recognition apps(7). Viewing angle, ambient lighting, and screen glare also varies. Screen size, resolution, brightness, and calibration may also vary across devices.

#### Optotype Sampling

In visual acuity tests, optotype sampling determines how subsequent optotypes are selected. Fixed sampling refers to predetermined sequences of optotypes e.g., letter sequence in ETDRS across lines. In adaptive sampling methods, sequences of letters or orientations of optotypes are randomized, thereby eliminating memorization bias. Different optotypes used to test patients may require demographic (e.g., symbols for young children) and regional (e.g., charts using letters in Roman and in Arabic letters), hence inducing another source of variability due to their differences in meaning, physical spatial contents. Orientation judgement paradigms using Landolt C or tumbling E optotypes avoid these intertest confounders as only optotype size and gap orientation is varied. In the context of intertest comparison, letter identification and orientation judgments undergo different neural computations, thus adding additional variability. Secondly, adaptive sampling uses various algorithm to select the next stimulus or set of stimuli based on previous responses. Additionally, some adaptive tasks do not have a fixed step size between varying optotype sizes, which allows finer resolution of acuity tests (6,7,10,22,23). The current gold-standard logMAR chart, such as ETDRS, includes 5-same sized-stimuli per line scaled at 0.10logMAR step size at a given distance using a 10 alternative forced choice task(24). The scoring is recommended such that all letters need to be identified correctly to receive the score, or stop at the line if this criterion is not met(24). Threshold or visual acuity is then determined as the last line that had no errors. However, some users may deploy a more lenient criterion, such as identifying 3 or 4 letters correctly before accepting the line, to account for various sources of noise(25). Scoring by letter-counting is another method (0.02 logMAR per correct letter) often applied. However, it discounts that the letter size within each line is the same. Additionally, it also dismissed the effect of a 60% correct criterion measures at a different point of the psychometric function than at 100% correct. Hence, these scoring methods are misleading and contributing to variance in VA measurements.

#### Response Input

The method by which an examinee’s responses are recorded is another source of variability. In conventional testings with letter-based charts (e.g., ETDRS, Snellen, and Bailey-Lovie) or symbol-based charts (e.g., LEA symbols), the examinee typically name each optotype out loud and the examiner judges whether the response is correct in order to continue to the next line or terminate the test. For orientation-based optotypes (e.g., Landolt C or Tumbling E), responses may be given verbally or by gesture. In each case, the examiner records the response manually or enters it into a computer system. An examiner’s judgement on scoring and error in calculation could introduce variability in VA measurements.

Alternatively, some VA assessments that enable examinee to enter their own responses using methods such as touchscreens(6,10,22,26), keyboards(16,23,27,28), or voice recognition(29). Each method has its own potential sources of error and may therefore affect the accuracy and precision of measurements in different ways.

#### Level of Supervision

Whether a VA assessment is supervised or not could be another variable that impact measurement accuracy and precision. In routine clinical testing, a trained technician is present in the room and administers the test. Their role may include providing instructions, ensuring consistent viewing distance, and scoring the results. The degree of examiner involvement, however, based on design of the assessment. Some tests heavily rely on the examiner, who is responsible for response input, optotype selection, and scoring. Others, such as automated tests with examinee self-input, require minimal involvement, and the examiner only serve to oversee the test and provide technical assistance as needed. Regardless of how involved the examiner is, it remains important to determine whether supervision is associated with accuracy and precision of VA testings.

#### Other factors: Viewing distance

Viewing distance is a further source of variation. The standard 6-meter (20 feet) distance approximates optical infinity, minimizing accommodation’s and pupil constriction’s contributions to threshold(5) whereas near- and intermediate VAs are typically measured at 40cm (16 in) and 67cm (26 in), respectively(30–32). Many tests use shorter distances such as 4 meters (13 ft) in ETDRS protocols(33), 1–2 meters on tablets and smartphones(34,35) for practical reasons. Distance can shift measured thresholds independent of device: one study attributed a systematic paper-versus-digital chart difference partly to differing test distances (3m or 10ft vs. 5m or 16ft) rather than display technology(36). Shorter distances can also alter crowding, since inter-optotype spacing does not always scale proportionally with letter size across displays, with downstream effects on both measured acuity and repeatability(37). Like procedure and scoring, distance is often confounded with modality, since remote and mobile tests skew toward shorter distances.

### Validation of visual acuity tests

Validation of VA tests is commonly established by repeatability and intertest agreement experiments. A repeatability experiment consists of comparison two-measurements of the same test. For intertest agreement experiment, a reference test is compared against a newly developed test to estimate its agreement. Note that during such experiment, no ground truth exists. Nevertheless, the comparison between two-tests has clinical utility in determining whether tests results are interchangeable. Consequently, without clear estimates of a VA-test’s repeatability and intertest-agreement, an interpretation of results within and between tests remains challenging and prevents meaningful judgement during clinical trials and in clinical practices. Agreement and repeatability can be evaluated using several statistical approaches, each with distinct limitations. The intraclass correlation coefficient (ICC) measures relative reliability but is influenced by between-participants variability and does not directly show the magnitude or direction of paired differences(38). The coefficient of variation (CoV), which expresses variability relative to the mean, does not directly assess paired agreement and can be difficult to interpret when logMAR values approach or cross zero. Bland-Altman analysis instead characterized paired differences using the mean bias and 95% limits of agreement (LoA)(39–41). For test-retest comparisons, the corresponding variability measure is expressed as the coefficient of repeatability (CoR), and the half width of 95% LoA (LoA half-width) for intertest comparisons. Accordingly, this review evaluated Bland-Altman derived mean bias, intertest LoA-half-width, and test-retest CoR.

### Prior literature

Prior literature has addressed some of the above-described heterogeneity in VA testings. Claessens et al. reported digital self-assessed VA accuracy via ranges of mean bias (range 0.08 to 0.10 logMAR) and 95% limits of agreement (0.08 to 0.47 logMAR)(42). Thirunavukkarasu et al. documented substantial inconsistency in how accuracy and reliability are reported across remotely self-administered VA tests(43). Van der Zee et al. investigated broader range of VA tests and reported the overall mean systematic bias of 0.03 logMAR and overall mean 95%LoA of ±0.20 logMAR in distance VA measured in adults(44). However, these previous reviews focused on digital and self-administered VA test in adults and did not separately characterize intertest and test-retest outcomes across testing characteristics.

The current study’s primary objective was therefore to evaluate the precision and accuracy of a visual acuity test’s repeatability and intertest agreement along four factors of variability: (1) test sites (clinic vs remote), (2) optotype sampling (fixed vs adaptive), (3) response input method (examiner vs self), and (4) level of supervision (supervised vs unsupervised). We hypothesized that VA tests precision will decreases with added sources of test variability. Moreover, based on prior meta-analyses(42,44), we hypothesized that accuracy outcomes are not significantly affected by increased sources of variability.

## Methods

### Search Strategy

A systematic review following the Preferred Reporting Items of Systematic Reviews and Meta-Analysis (PRISMA) 2020 guideline(45) was conducted using PubMed, Google Scholar, and Scopus. This study protocol was not pre-registered. Search terms and the full syntax can be found in the Supplementary Materials. The search filter included literature written in English and published between 1 January 1983 to 02 December 2024. Studies published before 1983 were excluded as it was before the publication of the seminal Bland-Altman analysis(46). Preprints without peer-review and conference abstracts were excluded from the review. Previously published reviews were also searched for relevant studies(42,43). Zotero (version 9.0.6) was used for extraction and screening of literature.

### Selection Process

The selection process involved two reviewers (C.C. and J.S.). Titles and abstracts were screened for relevance, and full text were reviewed for eligibility. C.C. performed the initial screening and selection. JS reviewed all selected papers and compared agreement with C.C. Disagreements were resolved through consensus. Systematic reviews, editorials, articles without accessible full-text, objective visual acuity assessments, and studies without Bland-Altman intertest or test-retest analyses were excluded. Detailed eligibility criteria are provided in the Supplementary Material. Figure 1 illustrated the screening and selection process based on PRIMSA guideline (Fig 1).

**Fig 1.**
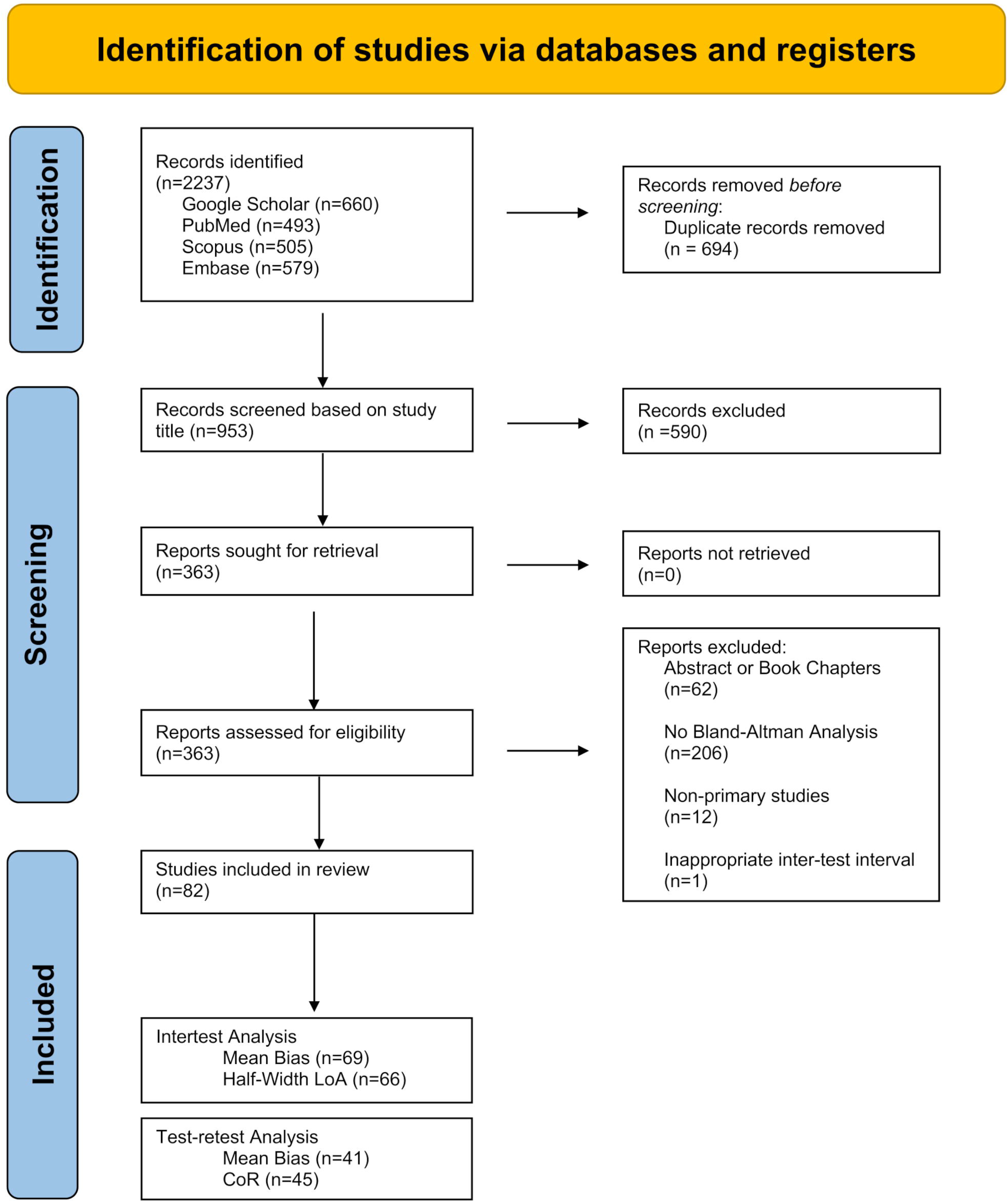
Preferred Reporting Items for Systematic Reviews and Meta-Analyses (PRISMA)(45) flowchart. Flow chart representing the study selection process. Total of 82 studies were included in this meta-analysis.

### Data Collection, Extraction and Synthesis

Reviewer C.C. extracted study characteristics and outcome data, including participant age, study design, index and reference test, testing parameters, mean bias, and upper and lower limits of agreement (ULoA and LLoA). When available, the standard deviation (SD) of paired differences and the standard of errors (SE) and 95% confidence interval (CI) for mean bias and LoA were also extracted. Mean bias represented the systematic difference between measurements and was standardized as index minus reference for intertest comparisons and first minus second measurements for test-retest comparisons.

All VA outcomes were expressed in logMAR, other VA units such as Snellen notation and decimal were converted to logMAR(47–50). LoA half-width for intertest comparison and CoR for test-retest comparisons were calculated as the distance between mean bias and ULoA (Fig 2). When mean bias was unavailable, the values were estimated as the average distance between ULoA and LLoA (estimated LoA = (ULoA-LLoA)/2. Graphically only data were approximated from the published figures (51–53). Reviewer J.S. audited the extraction and conversion of 20 randomly selected studies by comparing recorded data and the respective source articles. Discrepancies between reviewers were discussed and resolved over consensus.

**Fig 2.**
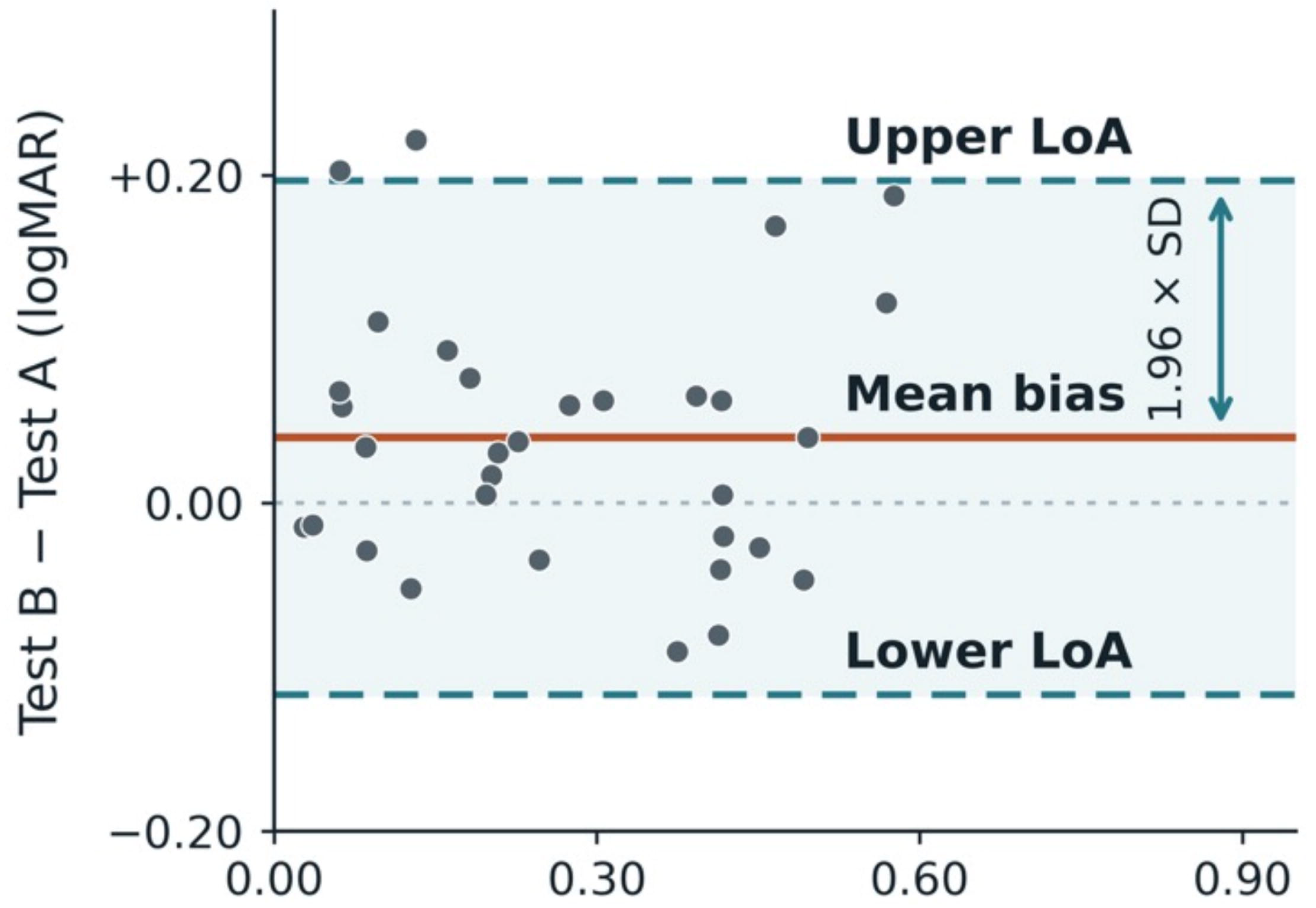
Visual representation of Bland-Altman mean bias and limits of agreement(39). 95% of paired differences for this hypothetical data set are expected between the upper and lower LoA indicated as blue dashed lines. For intertest agreement and for test-retest, the LoA half-width and coefficient of repeatability (CoR) are used to determine precision. Each dot represents one measurement pair. SD refers to standard deviation of paired difference. The black dashed line along zero depicts the ideal measurement difference for reference. The red line refers to the mean difference between measurements and is called mean bias. The y-axis shows the measurement differences while the x-axis shows the mean of the measurement. The image was generated via Image generated via ChatGPT, Version GPT-6.1 Sol using illustrative that assume normal differences with constant bias and variability.

When studies reported multiple eligible estimates, overall results were prioritized to avoid double counting. If tested for distance VA, ETDRS-based comparisons were selected when multiple reference tests were used. If tested for near VA, Rosenbaum-based comparison was chosen. If no overall estimate was available, right-eye data was preferred over separate monocular or binocular results.

### Primary Analysis – Testing Strategies

Each eligible study was classified according to their reported testing strategies based on testing sites, methodology of optotype sampling, methodology of response input, and level of supervision (Fig 3). The test site described *where* VA was assessed. Testing in a dedicated research or eye care facility was classified as “clinic,” whereas testing elsewhere, such as home, was classified as “remote.” Next, “Optotype sampling” describes *how* optotype sizes were generated and displayed. Since the optotype size sampling dictates what the actual optotype sizes in any given test are, it determines a VA task’s resolution. For example, VA tests with 0.10 logMAR size change per line on the ETDRS chart have a resolution of 0.10 logMAR and thus cannot measure any smaller changes. Tests using “fixed sampling” presented sizes in a predetermined sequence and terminated according to a pre-specified stopping rule, such as maximum number of errors at a given size. Examples included retro-illuminated ETDRS charts, Rosenbaum near acuity card, and digital charts with fixed increments(54–57). “Adaptive sampling” were defined as test that adjusted the optotype size according to preceding response to estimate VA threshold. Examples include staircase(12,16,21,35,58–61), adaptive charts(10), and Best-PEST(7,23,59) procedures. Response input described *who recorded* the examinee’s answers. Any study was classified as “examiner” when an examiner recorded a spoken or gestured response, and as “self” when an examinee self-entered a response using a touchscreen or keypad. Testing was classified as “supervised” when an examiner was present during the assessment and “unsupervised” when no examiner was present. Each eligible study was assigned to a testing-strategy group defined by its combination of test site, optotype sampling, response input, and supervision. Studies were excluded from testing-strategy analysis if the reported methods did not permit classification into all four categories.

**Fig 3.**
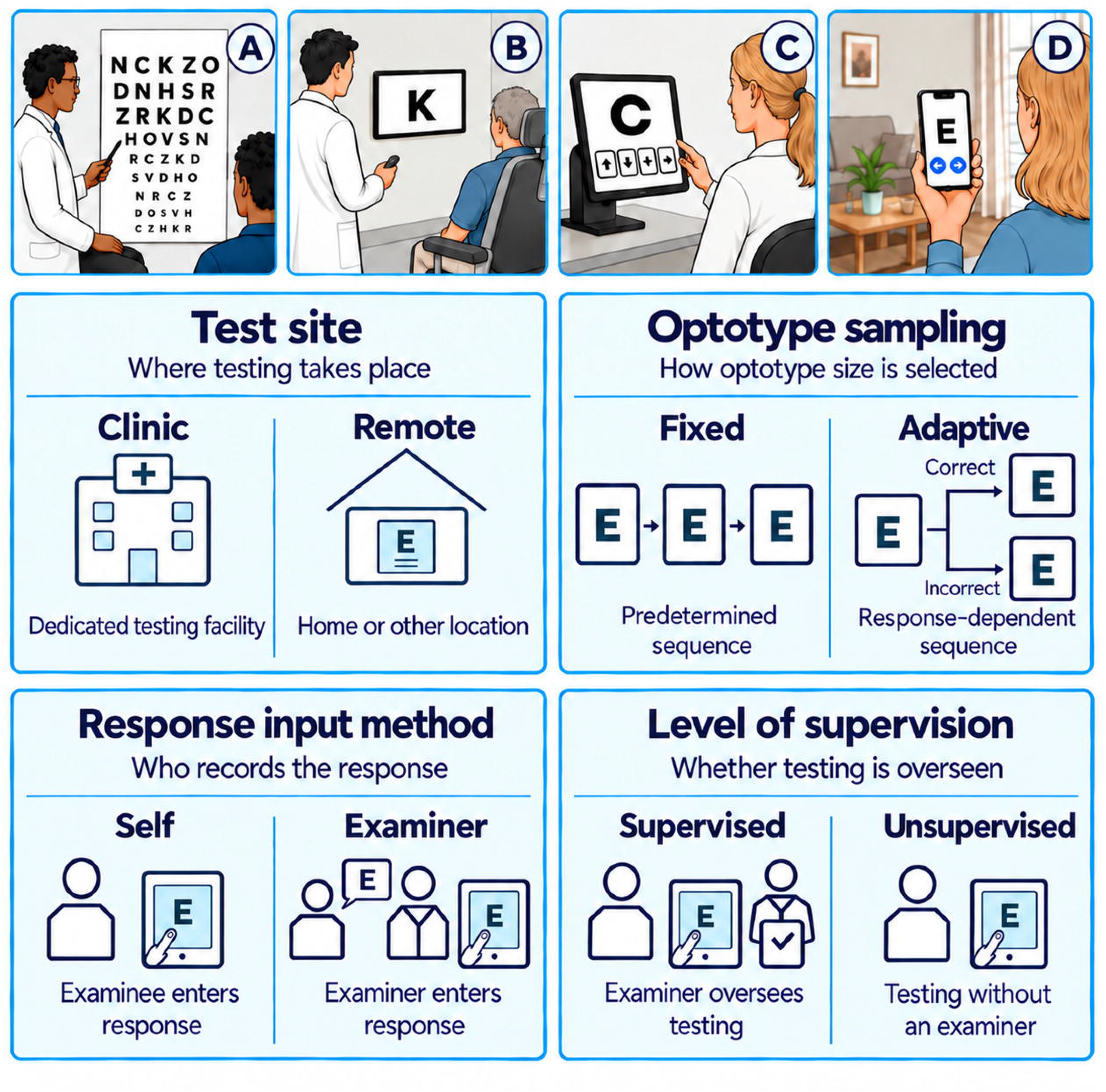
Schematic representation of different VA testing strategies. Top panel: represents the different strategies in assessing VA. It can be performed using a wall-mounted chart (A) or using a computerized system (B) under the presence of an examiner or self-administered using a tablet at a clinical facility (C) or using a personal mobile device at home (D). Bottom panels: Variability in testing methodology can be broken down into 4 different testing strategies: test site, optotype sampling, response input method, and level of supervision. Image generated via ChatGPT (OpenAI), Version GPT-6.1Sol.

### Subgroup Analysis – Patient Characteristics: Age and Clinical Status

Participants younger than 18 years were classified as children, and those aged 18 years or older were adults. Studies were assigned to the corresponding age group (Table 1). Estimates from mixed-age population without reporting results separately were excluded from age subgroup analysis. Similarly, effect estimates were classified by whether participants had ocular pathology. Studies that included populations with mixed or unclear pathology status were excluded from the pathology subgroup analysis.

**Table 1.**
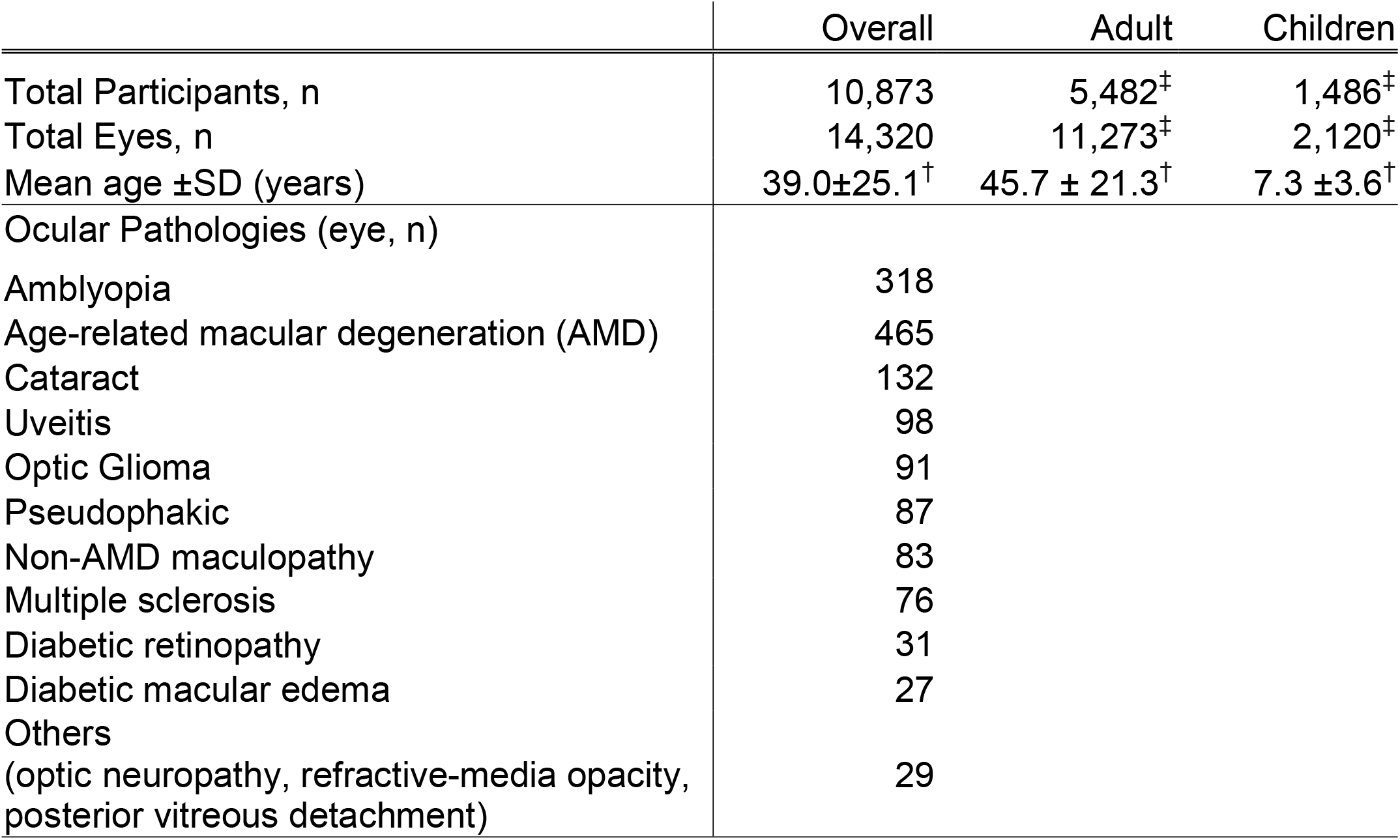
Summary of population characteristics. Numbers of participants and eyes, mean age (±SD), and ocular pathologies are summarized for all included studies and stratified by adults and pediatric populations. Data are limited to information reported and extractable from the included studies. ^†^Mean age estimated from publications that report both mean and SD. ^‡^Number of adult and children estimated from publications that reports adult and children as separate cohorts

### Statistical Analysis

Four outcomes were analyzed separately: intertest accuracy (mean bias), intertest precision (LoA half-width), test-retest accuracy (mean bias), and test-retest precision (CoR). All VA data were analyzed in or converted to logMAR units. Since all LoA half-width and CoR values included in the analysis were positive, their natural logarithms were analyzed to compare proportional differences between groups. Pooled estimates and 95% confidence intervals (CIs) for precision outcomes were converted back to original logMAR scale for presentation. Data were organized in Microsoft Excel and imported into R version 4.4.1 (R Foundation for Statistical Computing, Vienna, Austria) using with RStudio version 2026.07.01+147 (Posit Software, PBC, Boston, MA, USA) for all statistical analyses and graphical outputs. Three-level random-effects meta-regression models were fitted using the *metafor* package. These models accounted for multiple effect estimates from the same study and distinguished sampling error, variation among effects within studies, and variation between studies. Effect estimates, k, refer to data points in the meta-regression contributed by a study. For example, a study evaluating 2 different index VA tests across the same population will be contributing to 2 effect estimates. The within-study and between-study variation was estimated using restricted maximum likelihood (REML). An overall pooled estimate and 95% CI were calculated for each outcome. For each outcome, separate three-level meta-regression models were fitted for test sites, psychophysical procedure, response input, and supervision. An additional model compared the observed combination of these four testing strategies as distinct testing groups. Differences between testing groups were evaluated using study-clustered CR2 robust Wald tests with the HTZ small-sample adjustment using the *clubSandwish* package. When the test could not be calculated, the model-based moderator test was reported and identified as such.

### Study Risk of Bias Assessment

The Quality Assessment of Diagnostic Accuracy Studies 2 (QUADAS-2) tool was used to assess the risk of bias across four domains (patient selection, index test, reference standard, and flow and timing) and applicability concerns in each included study(62). Risk of bias was classified as either low, high, or unclear. Reviewer C.C. applied the QUADAS-2 signaling question to each study and discussed with reviewer J.S. The risk of bias judgements and summary graphs were visualized in traffic light plot generated via the Research Gold QUADAS-2 Tool(63).

## Result

A total of 2237 articles were identified through database searches. After removing duplicates and screening titles/abstracts, 363 full-text articles were assessed for eligibility, of which 78 studies met the inclusion criteria for meta-analysis. Of the 82 studies, 70 conducted intertest and 43 test-retest evaluations. After data extraction for the primary analysis, 69 studies were included for the intertest mean bias analysis, 66 for the intertest half-width LoA analysis, 41 for the test-retest mean bias analysis, and 45 for the CoR analysis (Fig 1).

### Study Characteristics

The 82 included studies comprised of a total of 10,873 participants and 14,320 eyes. Study population includes all ages (overall mean age 39.0 ±25.1; range of 2 - 93 years old). The most common ocular diagnosis was amblyopia, age-related macular degeneration, and cataracts (Table 1). The most used reference charts were ETDRS, followed by Snellen and Landolt C charts (Supplementary Material, Table S1). Selection of index tests varied in optotype used and testing strategies. Some uses retro-illuminated fixed chart, app-based fixed eye chart, app-based adaptive eye charts, or TV-based eye charts that utilize voice recognition to record responses(6,8,13,21,35,64–66). One effect estimate that utilized a non-optotypes-based testing was excluded from the analysis(56). ETDRS was the most used reference chart (effect estimates, k =145), followed by Snellen (effect estimates, k =39) and Landolt C charts (effect estimate, k =26). Smartphones, tablets, computers, and smart television are commonly used digital tools amongst examinee self-inputted VA tests.

The intertest mean bias and LoA half-width analyses included 69 and 66 studies, respectively, whereas test-retest mean bias and CoR analysis included 41 and 45 studies. Overall, 114 intertest and 101 test-retest effect estimates were included. Table 2 summarizes the numbers of studies (n) and effect estimates (k) within each testing strategy category. Studies evaluating multiple VA tests with different testing strategies could contribute multiple effect estimates and be counted in multiple categories.

**Table 2.** Summary of studies (n) and effect estimates (k) in each corresponding testing strategies. Majority of effect estimates conducted their VA test in clinics with responses recorded by the examiner under a supervised condition. Fixed and adaptive samplings had roughly balanced representation among the included studies.

| Testing Strategies | Category | Studies, n | Effects, k | Studies, n | Effects, k |
|---|---|---|---|---|---|
|  |  | Intertest |  | Test-retest |  |
| Test site | Clinic | 64 | 106 | 44 | 100 |
|  | Remote | 6 | 8 | 1 | 1 |
| Optotype Sampling | Fixed | 37 | 53 | 27 | 50 |
|  | Adaptive | 38 | 61 | 29 | 51 |
| Response-input | Examiner | 51 | 85 | 36 | 80 |
|  | Self | 21 | 29 | 15 | 21 |
| Supervision | Supervised | 64 | 107 | 44 | 100 |
|  | Unsupervised | 5 | 7 | 1 | 1 |

After reviewing the methodology of each included studies, we identified a total of 7 combinations across the 4 testing strategies. Combination and their corresponding studies can be found in Table 3 for intertest and Table 4 for test-retest.

**Table 3.**
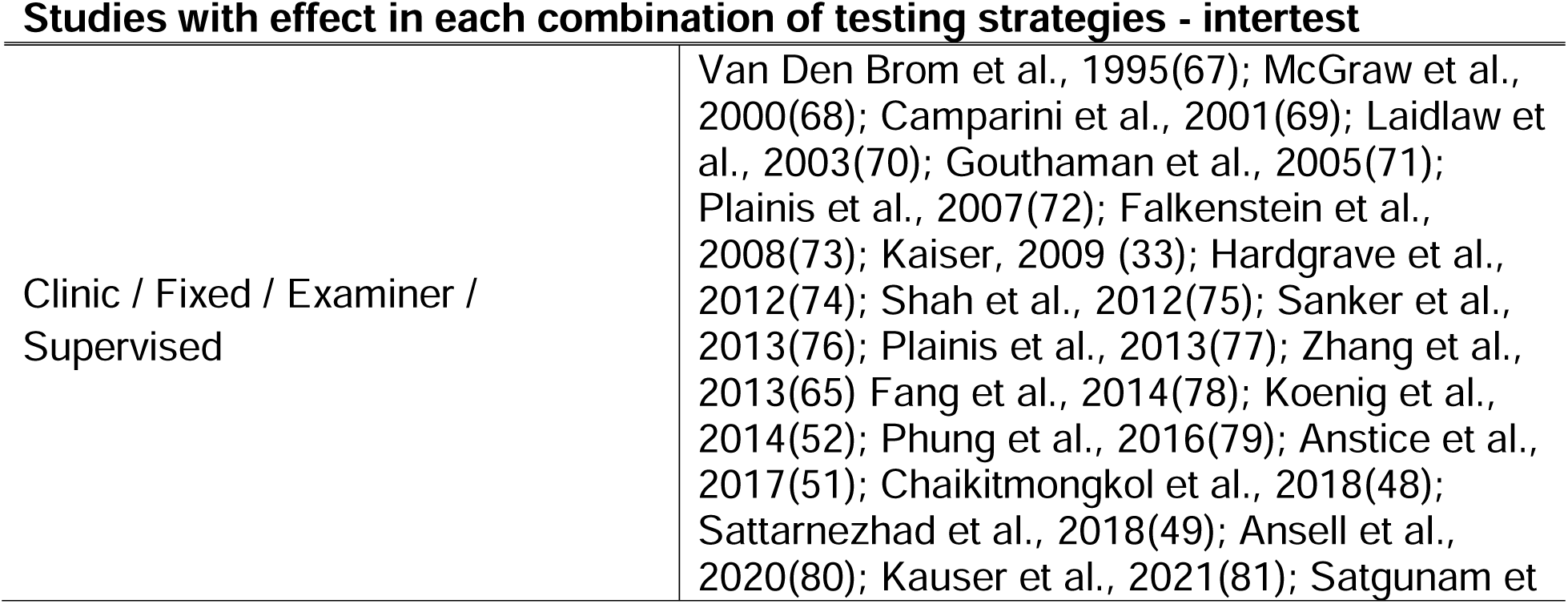
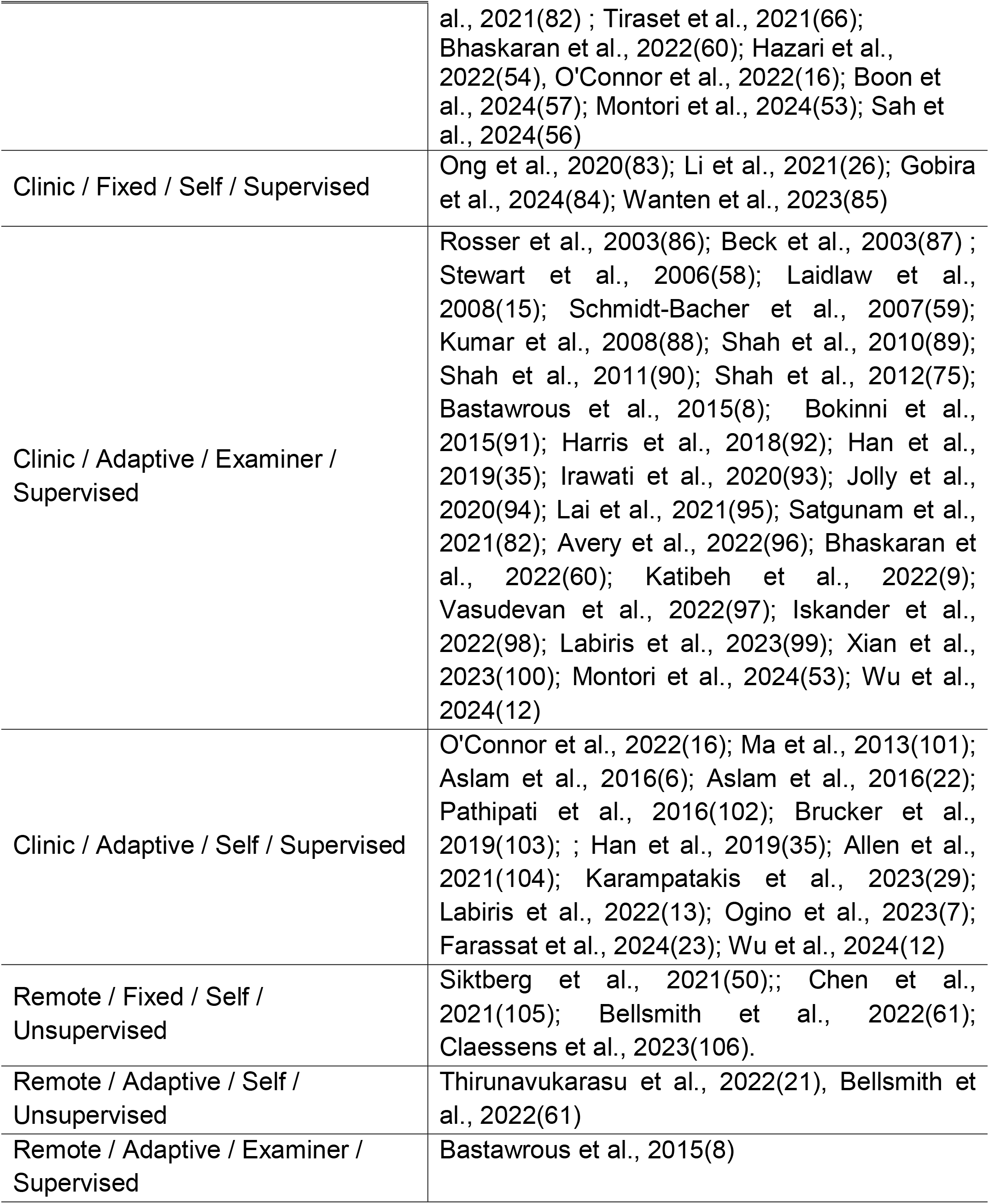
Summary of studies with intertest effect estimates fitting the criteria for each combination of testing strategies. Majority of the studies that performed intertest comparison were conducted under a controlled clinical facility.

| Studies with effect in each combination of testing strategies - intertest |  |
|---|---|
| Clinic / Fixed / Examiner / Supervised | Van Den Brom et al., 1995(67); McGraw et al., 2000(68); Camparini et al., 2001(69); Laidlaw et al., 2003(70); Gouthaman et al., 2005(71); Plainis et al., 2007(72); Falkenstein et al., 2008(73); Kaiser, 2009 (33); Hardgrave et al., 2012(74); Shah et al., 2012(75); Sanker et al., 2013(76); Plainis et al., 2013(77); Zhang et al., 2013(65) Fang et al., 2014(78); Koenig et al., 2014(52); Phung et al., 2016(79); Anstice et al., 2017(51); Chaikitmongkol et al., 2018(48); Sattarnezhad et al., 2018(49); Ansell et al., 2020(80); Kauser et al., 2021(81); Satgunam et al., 2021(82) ; Tiraset et al., 2021(66); Bhaskaran et al., 2022(60); Hazari et al., 2022(54), O'Connor et al., 2022(16); Boon et al., 2024(57); Montori et al., 2024(53); Sah et al., 2024(56) |
| Clinic / Fixed / Self / Supervised | Ong et al., 2020(83); Li et al., 2021(26); Gobira et al., 2024(84); Wanten et al., 2023(85) |
| Clinic / Adaptive / Examiner / Supervised | Rosser et al., 2003(86); Beck et al., 2003(87) ; Stewart et al., 2006(58); Laidlaw et al., 2008(15); Schmidt-Bacher et al., 2007(59); Kumar et al., 2008(88); Shah et al., 2010(89); Shah et al., 2011(90); Shah et al., 2012(75); Bastawrous et al., 2015(8); Bokinni et al., 2015(91); Harris et al., 2018(92); Han et al., 2019(35); Irawati et al., 2020(93); Jolly et al., 2020(94); Lai et al., 2021(95); Satgunam et al., 2021(82); Avery et al., 2022(96); Bhaskaran et al., 2022(60); Katibeh et al., 2022(9); Vasudevan et al., 2022(97); Iskander et al., 2022(98); Labiris et al., 2023(99); Xian et al., 2023(100); Montori et al., 2024(53); Wu et al., 2024(12) |
| Clinic / Adaptive / Self / Supervised | O'Connor et al., 2022(16); Ma et al., 2013(101); Aslam et al., 2016(6); Aslam et al., 2016(22); Pathipati et al., 2016(102); Brucker et al., 2019(103); ; Han et al., 2019(35); Allen et al., 2021(104); Karampatakis et al., 2023(29); Labiris et al., 2022(13); Ogino et al., 2023(7); Farassat et al., 2024(23); Wu et al., 2024(12) |
| Remote / Fixed / Self / Unsupervised | Siktberg et al., 2021(50);; Chen et al., 2021(105); Bellsmith et al., 2022(61); Claessens et al., 2023(106). |
| Remote / Adaptive / Self / Unsupervised | Thirunavukarasu et al., 2022(21), Bellsmith et al., 2022(61) |
| Remote / Adaptive / Examiner / Supervised | Bastawrous et al., 2015(8) |

**Table 4.** Summary of studies with test-retest effect estimates fitting the criteria for each combination of testing strategies. Majority of the studies that performed test-retest comparisons were conducted under a controlled clinical environment with supervision. Only one study tested remotely conducted a test-retest analysis.

| **Studies with effect in each combination of testing strategies – test-retest** |  |
|---|---|
| Clinic / Fixed / Examiner / Supervised | Van Den Brom et al., 1995(67); McGraw et al., 2000(68); Camparini et al., 2001(69); Beck et al., 2003(87) ; Rosser et al., 2003(86); Ruamviboonsuk et al., 2003(27); Laidlaw et al., 2003(70); Gouthaman et al., 2005(71); Chen et al., 2006(107); Stewart et al., 2006(58); Laidlaw et al., 2008(15); Lim et al., 2010(64); Shah et al., 2010(89); Shah et al., 2011(90); Hardgrave et al., 2012(74); Shah et al., 2012(75); Fang et al., 2014(78); Koenig et al., 2014(52); Bokinni et al., 2015(91); Jolly et al., 2020(94); O'Connor et al., 2022(16); Katibeh et al., 2022(9); Karampatakis et al., 2023(29); Iskander et al., 2022(98); Montori et al., 2024(53); Skerswetat et al., 2024(10). |
| Clinic / Fixed / Self / Supervised | Ruamviboonsuk et al., 2003(27); Li et al., 2021(26). |
| Clinic / Adaptive / Examiner / Supervised | Holmes et al., 2001(108); Beck et al., 2003(87) ; Cotter et al., 2003(109); Rosser et al., 2003(86); Stewart et al., 2006(58); Schmidt-Bacher et al., 2007(59); Kumar et al., 2008(88); Shah et al., 2011(90); Shah et al., 2012(75); Bokinni et al., 2015(91); Katibeh et al., 2022(9); Laidlaw et al., 2008(15); Jolly et al., 2020(94); Satgunam et al., 2021(82); 2022(98); Vasudevan et al., 2022(97); Montori et al., 2024(53); Wu et al., 2024(12); Han et al., 2019(35). |
| Clinic / Adaptive / Self / Supervised | Bach et al., 2006(28); Ma et al., 2013(101); Aslam et al., 2016(6); Aslam et al., 2016(22); Allen et al., 2021(104); O'Connor et al., 2022(16); Karampatakis et al., 2023(29); Farassat et al., 2024(23); Joseph et al., 2024(110); Wu et al., 2024(12); Skerswetat et al., 2024(10); Han et al., 2019(35). |
| Remote / Fixed / Self / Unsupervised | none |
| Remote / Adaptive / Self / Unsupervised | Thirunavukarasu et al., 2022(21). |
| Remote / Adaptive / Examiner / Supervised | none |

### Qualitative Assessment of selected studies

Using QUADAS-2 analysis(62), risk of bias was assessed across four domains: patient selection, index test, reference standard, and flow-and-timing (Fig 4). In the patient selection domain, 65% of the included study displayed high risk of bias. In the index test and reference standard domains, majority of the included studies have unclear risk of biases. Approximately half of the included studies (54%) displayed low risk of bias in the flow-and-timing domain with the remaining half split between high and unclear risk of bias.

**Fig 4.**
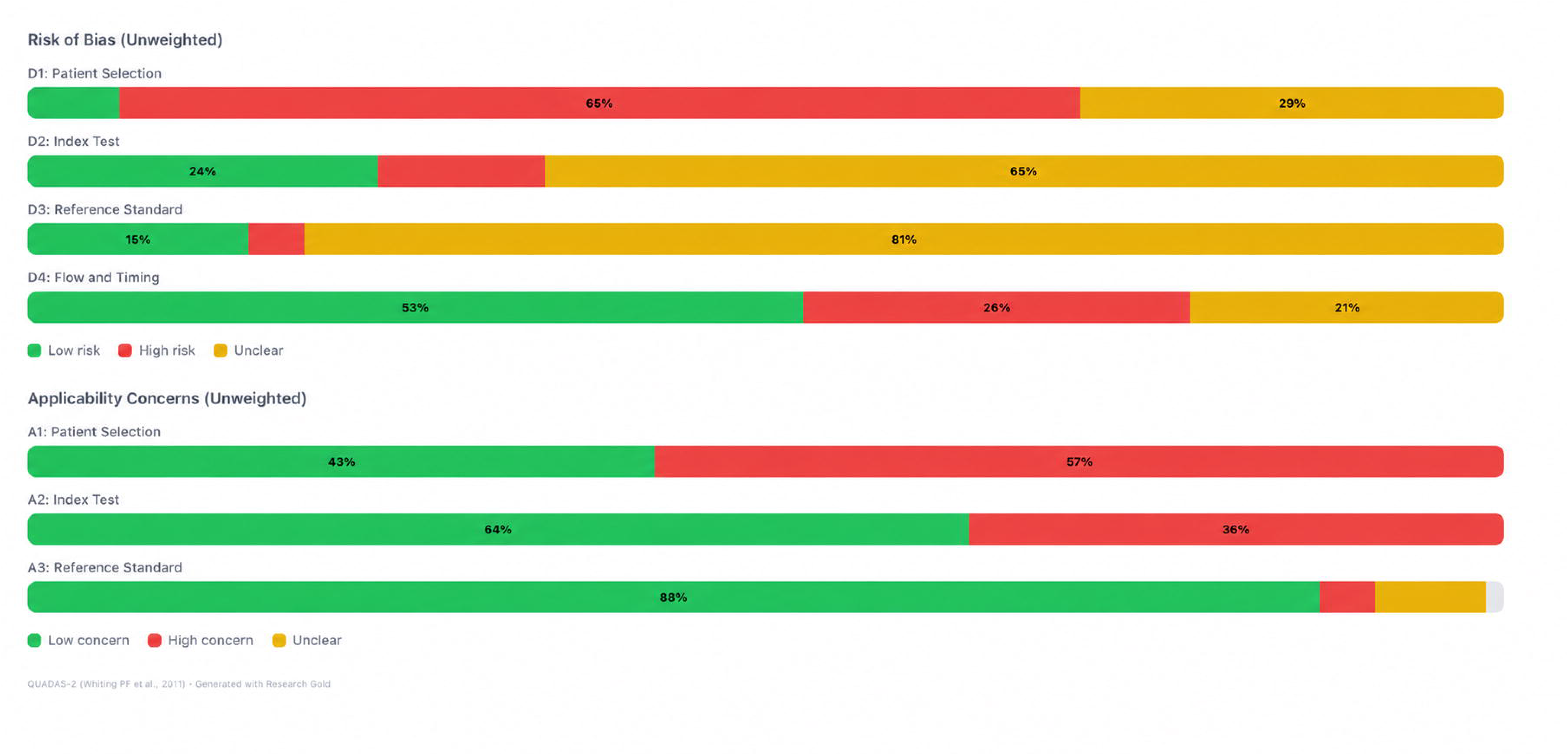
Summary of the QUADAS-2 risk of bias (RoB) and applicability concern analyses. Bars show the proportional of included studies rated as having low (green), high (red), or unclear (yellow) risk of bias or applicability concern within each domain. RoB was assessed across patient selection, index test, reference standard, and flow and timing. Applicability concerns were assessed for the first three domains. Font color for each percentage was edited from white to black using ChatGPT (OpenAI) Version GPT-6.1 Sol to improve legibility. Original image available in Supplement Figure S2.

Applicability concerns were evaluated across three domains: patient selection, index test, and reference standard. Low concern was observed in 41% of included studies for patient selection, 63% for the index test, and 87% for the reference standard. QUADAS-2 traffic light figure for individual studies can be found in Supplement Materials, Figure S1.

### Accuracy and Precision – Overall Result

Out of the 82 included studies, 69 studies and 114 effect estimates had extractable results for intertest accuracy (mean bias) analysis; 66 studies and 107 effect estimates were included in intertest precision (LoA half-width) analysis. Forty-one studies and 90 effect estimates were included in test-retest accuracy (mean bias) analysis; 45 studies and 101 effect estimates were included in test-retest precision (CoR) analysis (Table 5).

**Table 5.**
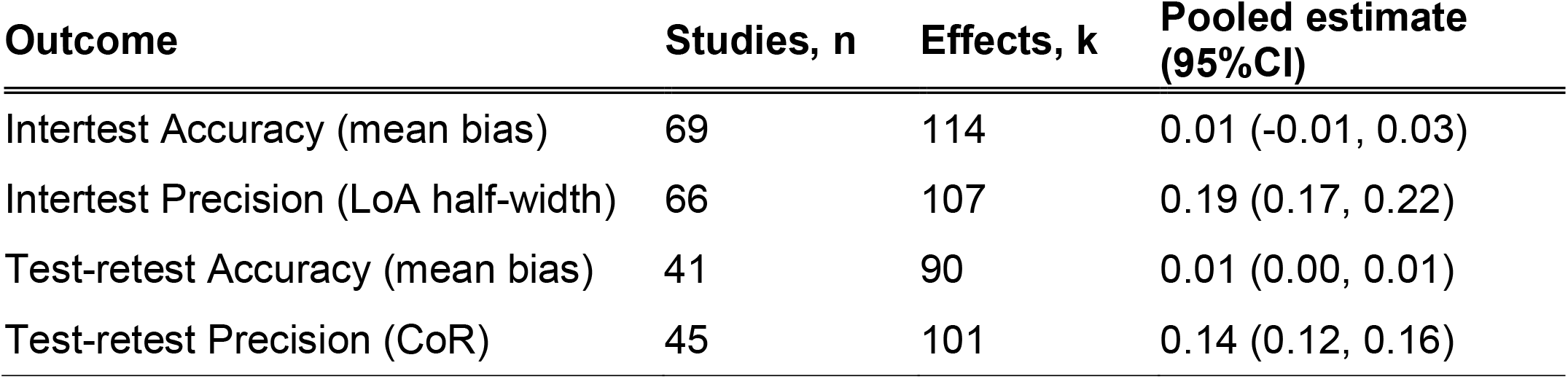
Overall pooled estimates for intertest and test-retest accuracy and precision. Table displayed number of included studies (n) and effects (k) with corresponding accuracy (mean bias) and precision (LoA half-width or CoR) metrics.

| **Outcome** | **Studies, n** | **Effects, k** | **Pooled estimate (95%CI)** |
|---|---|---|---|
| Intertest Accuracy (mean bias) | 69 | 114 | 0.01 (-0.01, 0.03) |
| Intertest Precision (LoA half-width) | 66 | 107 | 0.19 (0.17, 0.22) |
| Test-retest Accuracy (mean bias) | 41 | 90 | 0.01 (0.00, 0.01) |
| Test-retest Precision (CoR) | 45 | 101 | 0.14 (0.12, 0.16) |

**Table 6.** Summary of accuracy and precision benchmarks based on testing strategies. All meta-analytical results are shown in logMAR units and aided with their respective study sample size n. ^†^Result pooled from fewer than three effect estimates. ^‡^Testing strategy or combination of strategies ineligible for meta-analysis due to having with < 1 effect estimates.

| Testing Strategy |  | Intertest Agreement |  | Test-retest Repeatability |  |
|---|---|---|---|---|---|
|  |  | Accuracy (mean bias) | Precision (LoA half-width) | Accuracy (mean bias) | Precision (CoR) |
| Test site | Clinic | 0.01 [n=64] | 0.19 [61] | 0.01 [n=40] | 0.14 [n=44] |
|  | Remote | 0.01 [n=6] | 0.27 [n=6] | N/A<sup>‡</sup> [n=1] | N/A<sup>‡</sup> [n=1] |
| Optotype Sampling | Fixed | 0.02 [n=37] | 0.18 [n=37] | 0.01 [n=23] | 0.13 [n=27] |
|  | Adaptive | 0.00 [n=38] | 0.20 [n=35] | 0.01 [n=28] | 0.15 [n=29] |
| Response-input method | Examiner | 0.01 [n=51] | 0.19 [n=48] | 0.01 [n=32] | 0.13 [n=36] |
|  | Self | 0.00 [n=21] | 0.19 [n=21] | 0.00 [n=15] | 0.18 [n=15] |
| Level of supervision | Supervised | 0.01 [n=64] | 0.19 [n=61] | 0.01 [n=40] | 0.14 [n=44] |
|  | Unsupervised | -0.01 [n=5] | 0.26 [n=5] | N/A<sup>‡</sup> [n=1] | N/A<sup>‡</sup> [n=1] |
| **Overall** |  | **0.01** | **0.19** | **0.01** | **0.14** |
| Clinic / Fixed / Examiner / Supervised |  | 0.02 [n=30] | 0.18 [n=30] | 0.01[n=22] | 0.13 [n=26] |
| Clinic / Fixed / Self / Supervised |  | 0.01 [n=3] | 0.12 [n=3] | -0.01 [n=2] | 0.14 [n=2] |
| Clinic / Adaptive / Examiner / Supervised |  | 0.00 [n=25] | 0.20 [n=22] | 0.01 [n=17] | 0.14[n=18] |
| Clinic / Adaptive / Self / Supervised |  | 0.00 [n=13] | 0.20 [n=13] | 0.00 [n=12] | 0.19 [n=12] |
| Remote / Fixed / Self / Unsupervised |  | 0.00 [n=4] | 0.27 [n=4] | N/A<sup>‡</sup> [n=0] | N/A<sup>‡</sup> [n=0] |
| Remote / Adaptive / Self / Unsupervised |  | -0.03<sup>†</sup> [n=2] | 0.25<sup>†</sup> [n=2] | N/A<sup>‡</sup> [n=1] | N/A<sup>‡</sup> [n=1] |
| Remote / Adaptive / Examiner / Supervised |  | N/A<sup>‡</sup> [n=0] | N/A<sup>‡</sup> [n=0] | N/A<sup>‡</sup> [n=0] | N/A<sup>‡</sup> [n=0] |

### Intertest Accuracy- mean bias

The overall pooled intertest accuracy (mean bias) across 69 studies and 114 effect estimates was 0.01 logMAR [95%CI −0.01, 0.03], ranging from −0.22 to 0.24 logMAR (Supplementary Material Figure S3). Within each testing strategies, there was no significant difference in intertest accuracy between each paired-wise comparison (clinic vs remote; fixed vs adaptive; examiner vs self; supervised vs unsupervised). There was a trend toward improved accuracy in adaptive versus fixed psychophysical procedure, mean bias 0.00 logMAR [−0.01, 0.02] vs 0.02 logMAR [0.00, 0.05], but the difference was not statistically significant (p=0.060). Detailed result table can be found in Supplemental Material Table S2. When comparing intertest accuracy across different combination of testing strategies, there was no significant difference across groups (p=0.134). “Remote / Adaptive / Examiner / Supervised” was represented by one single study and effect, hence it was omitted in the comparison (Fig 5).

**Fig 5.**
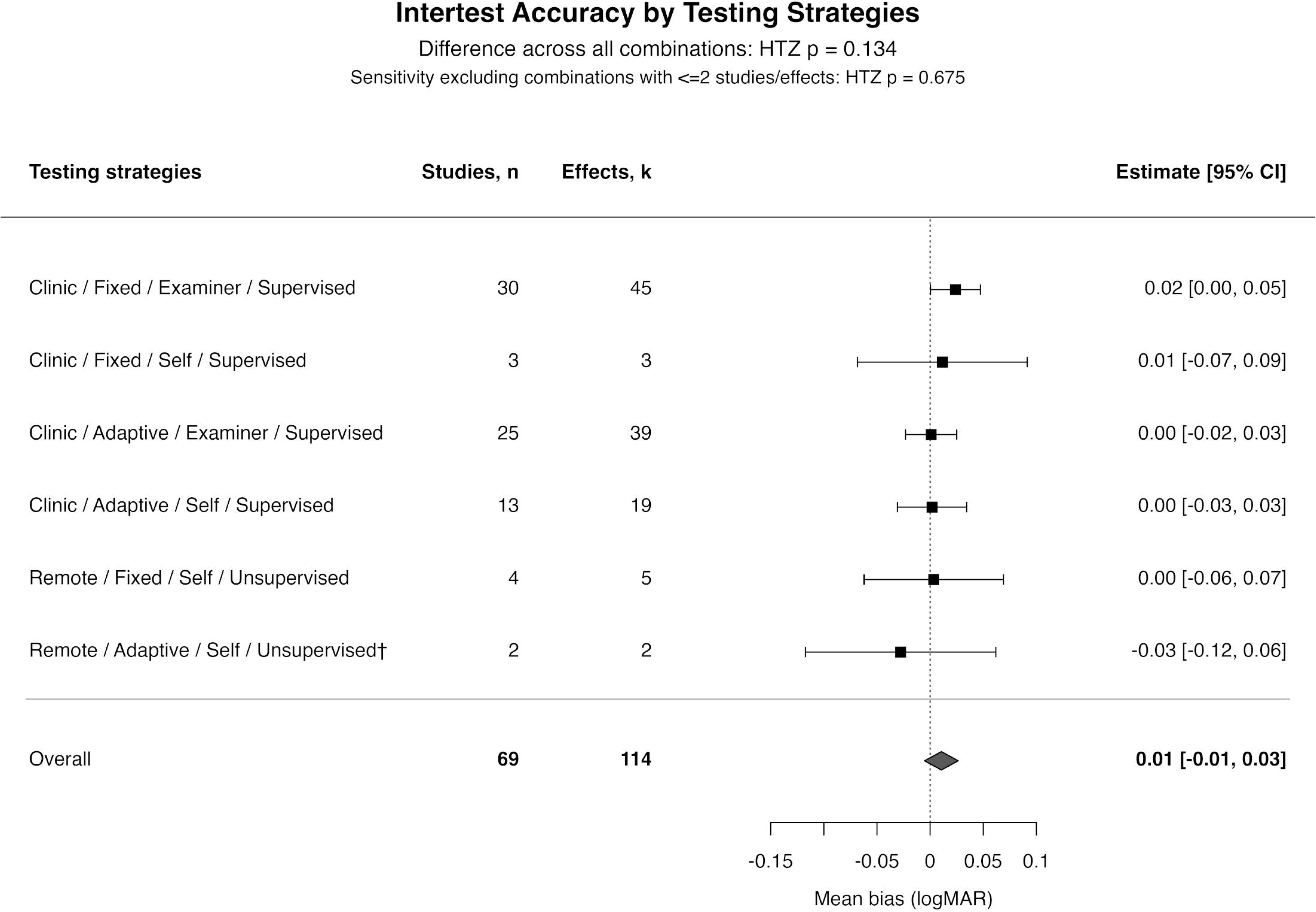
Summarized intertest mean bias forest plot stratified by testing strategies. Intertest accuracies were near 0.00 logMAR across combination of testing strategies. Squares and horizontal error bars represent pooled mean biases and 95% CIs. The diamond represents the overall pooled estimate. The vertical dotted line represents no systematic difference (0.00 logMAR). All data are represented as logMAR values.

### Intertest Precision – LoA half-width

Pooled intertest precision across 66 studies and 107 effect estimates was 0.19 logMAR [0.17, 0.22], ranging from 0.04 to 0.80 logMAR (Supplementary Material Figure S4). In comparison of intertest precision (LoA half-width) within each paired-wise comparison of each testing strategy, there was a significant difference between LoA half-width of VA measured under a controlled clinical facility versus remotely (0.19 logMAR [0.17, 0.21] for clinic vs 0.27 [0.20, 0.38] for remote, p=0.03). There was no significant difference observed in any other paired-wise comparison within each testing strategy. Detailed result table can be found in Supplemental Material Table S3. When comparing intertest precision across different combinations of testing strategies, there was no significant difference across groups (p=0.452). There was a trend toward increasing variability with less examiner involvement and when conducted under a remote facility (Fig 6).

**Fig 6.**
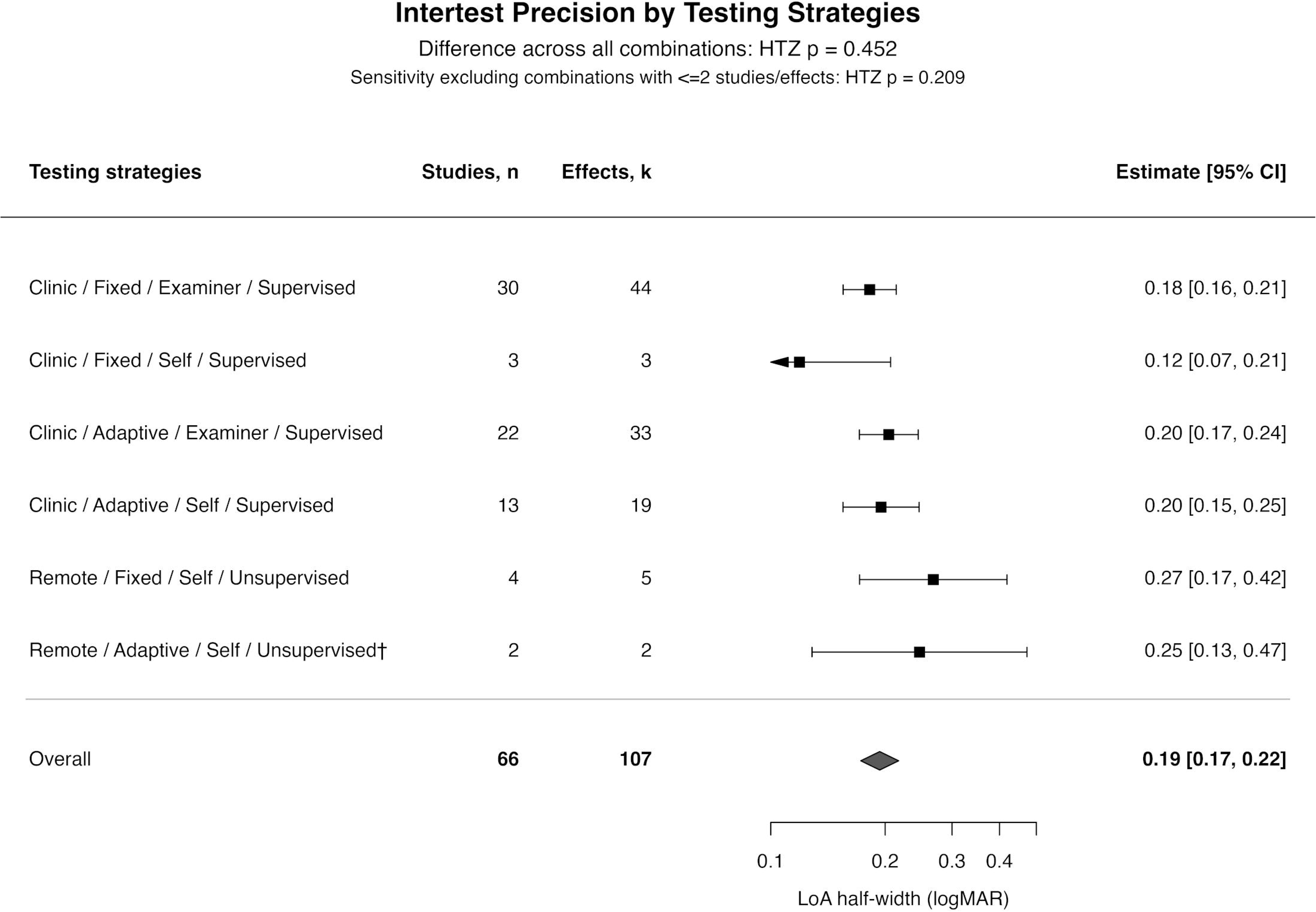
Summarized intertest LoA half-width forest plot stratified by combination of testing strategies. Overall results showed substantial intertest variability with LoA half-width of 0.19logMAR, and increasingly less precise with remote test sites. Squares and horizontal error bars represent subgroup estimates and their 95% CIs. The diamond represents the overall pooled estimate. All data are represented as logMAR values. ^†^The following combinations have two or fewer studies/effects.

### Test-retest Accuracy – mean bias

The overall pooled test-retest accuracy across 41 studies and 107 effect estimates was 0.01 logMAR [0.00, 0.01], ranging from −0.05 to 0.08 logMAR (Supplementary Material Figure S5). Among the four testing strategies, subgroup comparisons were feasible only for optotype sampling and response-input method. There was a statistically significant difference in test-retest accuracy between VA with examinee self-inputted response versus examiner-inputted (p=0.04), but the difference was not clinically significant (0.00 logMAR vs 0.01 logMAR for self vs examiner, respectively). There was no significant difference between utilization of fixed versus adaptive sampling (p=0.09). Test site and level of supervision were not compared because the remote and unsupervised subgroups each contained only one effect estimate. Detailed result table can be found in Supplemental Material Table S4. When comparing test-retest accuracy across different combinations of testing strategies, there was no significant difference across groups (p=0.426). “Remote / Adaptive / Self / Unsupervised” was represented by one single study and effect, hence it was omitted in the comparison (Fig 7).

**Fig 7.**
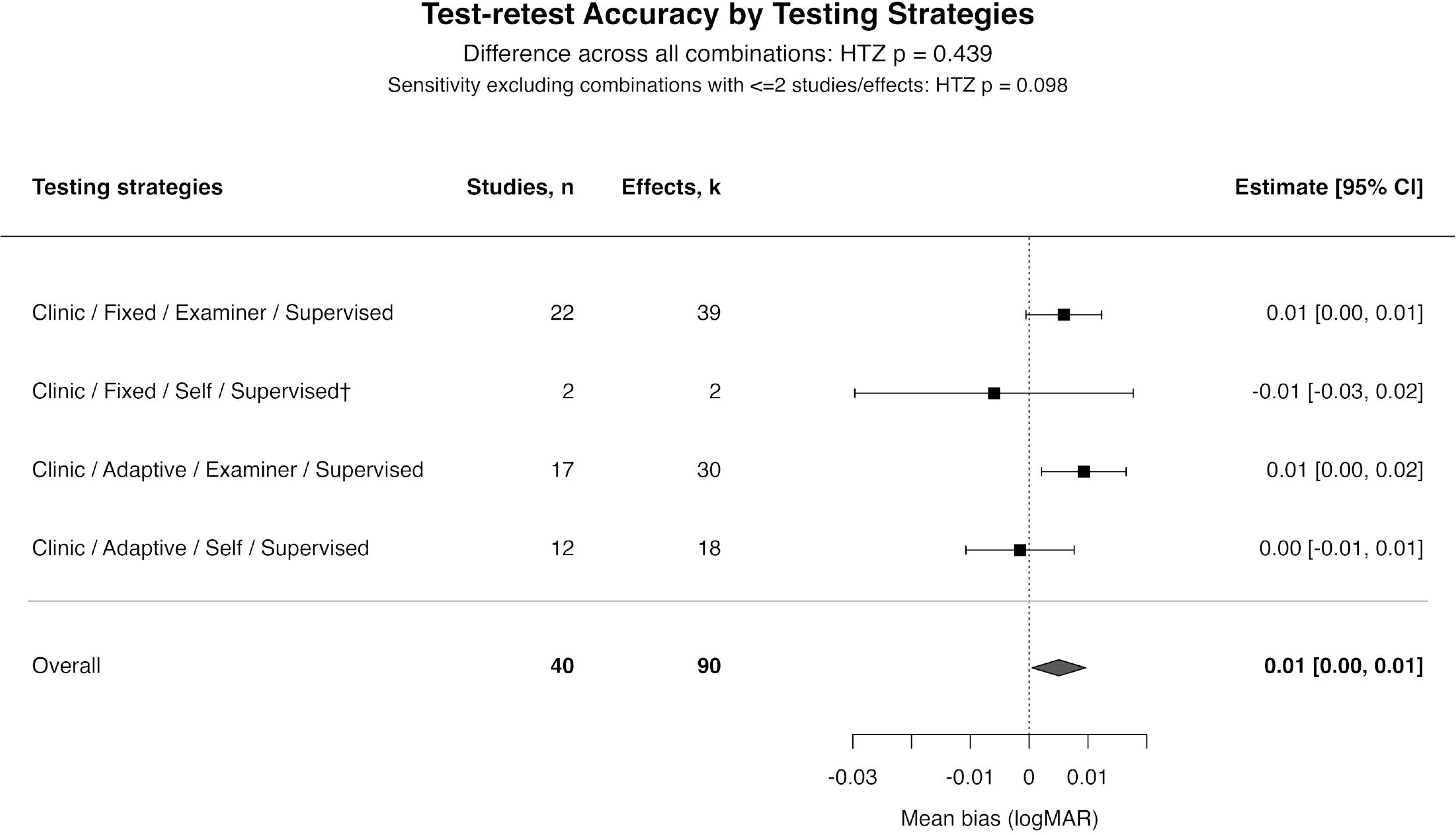
Summarized test-retest mean bias forest plot stratified by testing modalities. Overall demonstrated good accuracy across combination of testing strategies with overall mean bias of 0.01 logMAR. Squares and horizontal line represent pooled mean bias and 95% CI. The diamond represents the overall pooled estimates. The vertical dotted line represents no systematic difference (0.00 logMAR). All data are represented as logMAR value. Testing strategies including only one study was omitted in the forest plot; data remain in the overall estimate and omnibus test. ^†^The combinations have two or fewer studies/effects

### Test-retest Precision – Coefficient of Repeatability (CoR)

The overall pooled test-retest precision (CoR) across 45 studies and 101 effect estimates was 0.14 logMAR [0.12, 0.16] ranging from 0.02 to 0.39 logMAR (Supplementary Material Figure S6). There was a statistically significant difference in test-retest precision between VA with examinee self-inputted response (0.13 logMAR [0.11, 0.15]) versus examiner-inputted (0.18 logMAR. [0.14, 0.23]; p=0.030). There was no significant difference between utilization of fixed versus adaptive sampling (p=0.098). Test site and level of supervision were not compared because the remote and unsupervised subgroups each contained only one effect estimate. Detailed result table can be found in Supplemental Material Table S5. When comparing test-retest precision across different combination of testing strategies, there was no significant difference across groups (p=0.426). “Remote / Adaptive / Self / Unsupervised” was represented by one single study and effect, hence it was omitted in the comparison (Fig 8).

**Fig 8.**
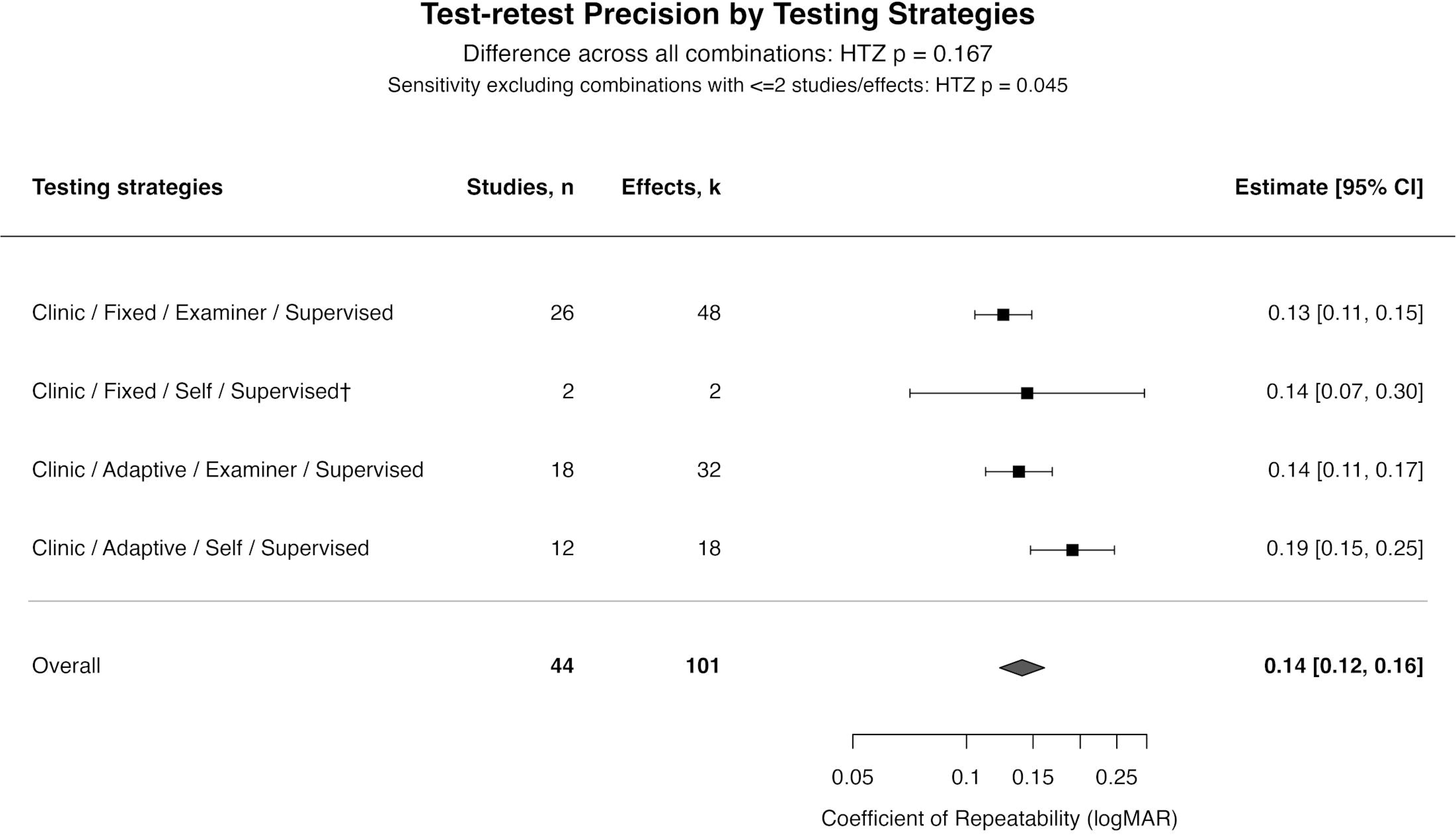
Summarized test-retest CoR Forest plot stratified by testing strategies. Overall wide CoR of 0.14 logMAR. Squares and horizontal line represent subgroups estimates and 95% CIs, and diamond represents the overall pooled estimate. All data are represented as logMAR value. Testing strategies including only one study was omitted in the forest plot; data remain in the overall estimate and omnibus test. ^†^The combinations have two or fewer studies/effects.

### Secondary Analysis – Age and Visual Acuity

Since VA is affected by age(111), we also investigated whether accuracy and precision differ between children and adults. Studies were stratified by reported participant age into adult (≥18 years), children (age<18 years). The pooled intertest accuracy (mean bias) was significantly different between adult 0.02 logMAR [0.01, 0.04] and children - 0.04 logMAR [−0.07, −0.01) in children (p=0.008). The corresponding intertest precision (LoA half-width) was not significantly different between the adult 0.19 logMAR [0.17, 0.22] and children 0.20 logMAR [0.16, 0.26), p=0.625. Test-retest accuracy (mean bias) did not differ significantly between the adult and children (p=0.734). The corresponding precision (CoR) was 0.13 logMAR [0.11, 0.16] in adult and 0.15 logMAR [0.12, 0.20] in children with no significant difference between the groups (p=0.471). Forest plots can be found in the Supplement Material, Figure S7.

### Secondary Analysis – Pathologies and Visual Acuity

Studies were categorized into “without pathology” or “with pathology” based on reported ocular health status. The intertest accuracy (mean bias) was 0.00 logMAR [−0.03, 0.03] for “without pathology” and −0.01 logMAR [−0.04, 0.02] for “with pathology” subgroups (p=0.671). The intertest precision (LoA half-width) was 0.17 logMAR [0.14, 0.20] for “without pathology” and 0.18 logMAR [0.15, 0.22] for “with pathology” subgroup (p=0.467).

Test-retest accuracy (mean bias) was 0.01 logMAR [0.00, 0.02] for “without pathology” and 0.00 logMAR [0.00, 0.01] for “with pathology” subgroup (p=0.405). Test-retest precision (CoR) weas 0.14 logMAR (0.11, 0.18) for “without pathology” and 0.13 logMAR [0.10, 0.16) for “without pathology” subgroups (p=0.387). There was no significant difference in intertest or test-test accuracy and precision across ocular pathology subgroup analysis. Forest plot based on ocular health status subgroups can be found in the Supplement Material, Figure S8.

## Discussion

Prior investigations have proposed benchmarks for interpreting variability in VA measurements. Siderov and Tiu suggested that a difference of at least 0.15 logMAR was needed to confidently identify a real change under routine clinical conditions(112). Rosser et al. found that changes of at least 0.20 logMAR could reliability distinguished from no change using the ETDRS chart(113). More recently, Van der Zee et al. reported an overall mean random error of 0.20 logMAR across intertest and test-retest comparisons and proposed this values a provisional frame of reference(44). Building on this work, our meta-analysis provides separate pooled estimates of intertest and test-retest variability as each has distinct clinical implications. Intertest variability addresses interchangeability between different tests, whereas test-retest variability helps distinguish true change in visual function from measurement error of the test itself when the same test was used repeatedly during longitudinal follow up. The 0.19logMAR in overall intertest precision implied that switching tests can introduce substantial variation and even more pronounced with assessments conducted in non-clinical test sites. These findings highlight the caution needed when integrating new VA tests into clinical practice or substituting them for established assessments. Although eye care facilities often have established protocol for routine VA testing, maintaining consistent testing conditions is particularly important when these assessments are extended beyond professional eye care settings. Examples include emergency departments, inpatient facilities and telehealth consultations, where rapid VA assessment is needed but testing environment may be difficult to standardize and testing instructions may be difficult to verify. Additionally, the 0.14 logMAR in test-retest precision reflects that small changes on repeated testing may be difficult to distinguish from expected measurement variabilities. Although the CoR of 0.14 logMAR fell within the 0.20 logMAR proposed by Rosser et al. and Van Der Zee et al., it remained greater than one ETDRS line (0.10 logMAR), suggesting that changes of this magnitude or smaller may still be obscured by measurement noise. Further reduction in test-retest variability could improve sensitivity to subtle longitudinal changes in VA.

### Intertest agreement

In this meta-analysis, systematic accuracy and precisions of validated visual acuity assessments were quantified across testing strategies and participants’ characteristics. The overall index VA tests demonstrated pooled systematic intertest mean bias very close to 0.00 logMAR, indicating the VA measured with index tests provided very similar results as the reference test. However, with the pooled systematic LoA half-width of 0.19 logMAR, it indicates we are expected to see variation of 0.19 logMAR (approximately 2 ETDRS lines) when comparing results from different tests. When different charts are used at follow-up visits, a change of less than 0.19 logMAR may clinically indicate a change in vision, but mathematically, may be difficult to distinguish from the expected variability between the tests.

When dissecting into different testing strategies, we observed a statistically significantly wider (p=0.030) intertest LoA half-width between VA conducted in non-clinical test site (“remote”) and those conducted in a controlled clinical test site (“clinic”). This observation was also present when comparing intertest precision across different combination of testing strategies. Though the comparisons did not reach a statistically significant threshold when evaluating testing strategies among different combinations, there was a trend toward greater variability (LoA half-width 0.27 and 0.25 logMAR, Figure 5) in testing strategies involving remote testings. One consideration for the observed difference in variability was the imbalance of sample size. The remote subgroup included only 6 studies and 8 effect estimates, compared with 61 studies and 99 effect estimates in the clinic subgroup, making the remote pooled estimate more sensitive to individual studies. Another aspect to consider was that remote testings often coincided with unsupervised and examinee self-inputted responses, making the contribution of these characteristics difficult to differentiate. However, on the paired-wise comparison within each testing strategy, LoA half-width did not differ significantly by input method (examiner vs self; p=0.925) and level of supervision (supervised vs unsupervised; p=0.108). One “Remote / Adaptive / Examiner / Supervised” combination had a LoA half-width of 0.48 logMAR, which is more than twice of the overall pooled value of 0.19 logMAR. This combination was omitted from the combination level analysis was it was represented by a single effect estimate(8). However, it could still contribute to the variability in overall pooled LoA half-width in the “remote” subgroup. Regardless, it highlights the substantial variability in remote testing can still occur even when conducted with the examiner input and under supervision.

In intertest agreement, testing conditions with increase sources of variability were associated with widen LoA half-width across VA tests, while differences in bias were marginal. These findings suggest that precision is more susceptible than accuracy to potential sources of variability in VA testing strategies and highlight that VA measurements can be accurate on average yet imprecise at the individual level.

### Repeatability

We also observed a similar result pattern for test-retest comparisons. The pooled systematic test-retest mean bias was very close to 0.00 logMAR, indicating no overall learning or fatigue effects that would yield a better or worse results upon repeated testing. However, the lack of precision was evident. The systematic pooled test-retest CoR of 0.14 logMAR indicates if the same test were to be repeated, a difference that’s smaller than 0.14 logMAR would be mathematically considered as the measurement variability within the test, not a true meaningful change. While the solution for intertest variability would be addressed by using the same eye chart and testing strategies when following up with patients, the lack of precision in test-retest poses a greater limitation in advancing assessments with wide test-retest CoR for clinical use, especially when monitoring change in subtle VA is crucial in evaluating treatment outcome and influencing clinical decision making.

In evaluation of test-retest CoR within each testing strategies, we found a significant difference in CoR between tests that utilized examiner-input versus examinee self-input (p=0.030). The CoR for self-inputted tests to be wider than those of examiner-inputted (0.18 vs 0.13 logMAR; p=0.030), but the difference of 0.05 logMAR may not be clinically significant. The increased variability in self-inputted tests could be explained by the differences in how examinee self-inputted their responses. Some utilized keypads labeled with corresponding orientation of the optotype(23,27,28) or corresponding Kay Pictures(16). Others utilized touchscreen devices(6,10,22,26) or voice recognitional system for response input(29). In addition to the methodology of how the responses were recorded, another variable worth mentioning is whether different psychophysical procedures could influence the variability observed in self-inputted VA tests. In fixed procedure, the scoring rules were line-by-line or letter-by-letter with relatively coarse increment. In adaptive procedure, optotype sizes were adjusted based on preceding responses and enable finer increments of VA measurement. Finer scoring in adaptive procedure may capture smaller differences in response that would be rounded to the same acuity value with line-based scoring. Thus, increasing the observed measurement variability. This is consistent with the findings in our analysis, where adaptive psychophysical procedure has wider CoR than fixed, but the differences did not reach a significant level. (0.15 vs 0.13 logMAR; p=0.098).

Regardless, within self-inputted VA tests, 13 studies utilized adaptive and 2 studies utilized fixed psychophysical procedure. The predominant representation of adaptive procedure in self-inputted test may contribute to the variability in self-inputted VA tests. In evaluation of CoR among different combination of testing strategies, we observed similar pattern as described above. Test-retest CoR was the least precise in the combination that included adaptive procedure and examinee self-inputted testing (CoR 0.19 logMAR) compared to those of fixed procedures and examiner-inputted testings (CoR 0.13 and 0.14 logMAR), though again, the differences were not significant. Overall, we observed decrease in precision in testings that involved adaptive psychophysical procedure and examinee self-inputted testing strategies.

### The effect of test resolution in optotype sampling on precision during repeatability experiments

Of the 38 studies that involved adaptive sampling, majority of the studies (n=32) relied on ETDRS as prototype of their chart design, which confined their step-size resolution to 0.10 logMAR. Similarly, most testing with fixed procedure (i.e., either fixed chart-based or fixed adaptive tests) followed the design of the ETDRS chart. Furthermore, some adaptive sampling methods may have additional free parameters (e.g. slope, lapse rate, differing Bayesian prior settings, etc.) and thus have more degree of freedom for test variability. Consequently, future studies should investigate the relationship between test resolution and different of parameters in adaptive protocol.

#### Influence of age and ocular pathology on test accuracy and precision

Beyond testing strategies, we also evaluated accuracy and precision on *who* were tested on the VA tests. Previous work(44) excluded individuals younger than 18 years, in part because the use of age-appropriate charts and optotypes were expected to introduce additional heterogeneity. Most included studies involving children used charts or optotypes also commonly used in adults, including ETDRS, Snellen, or Landolt C. One study used Baile-Lovie acuity chart(76), and another study used Kay Pictures(75). In our meta-analysis, we found a statistically significant difference in intertest accuracy (mean bias) of VA measured between adults and children (0.02 vs −0.04 logMAR; p=0.008). The negative mean bias indicates that index tests yielded lower logMAR values, corresponding to better VA, than reference tests. In another word, index tests measured better VA than references tests by 0.04 logMAR in children and worse VA than references by 0.02 logMAR in adults. Although intertest mean bias differed significantly between children and adults, the absolute between-group difference was small at 0.06 logMAR, suggesting the difference may have limited clinical relevance. No significant difference between children and adults were detected in intertest precision, test-retest accuracy, or test-retest precision. In both age groups, LoA-half width and CoR estimates were similar to the systematic pooled estimates. These findings provide no clear evidence that pediatric inclusion or use of age-appropriate testing reduced VA agreement or repeatability.

The presence of ocular pathologies may also influence accuracy and precision of VA measurements. Amblyopia and age-related macular degeneration (AMD) were the most represented conditions in this dataset. These conditions could theoretically affect performance through unstable fixation, impaired contrast perception, or fluctuating visual function(114). Nevertheless, test-retest CoR did not differ significantly between those without versus those with ocular pathologies (0.14 vs 0.13 logMAR; p=0.387). Similarly, there was no significant difference in intertest LoA half-widths between either pathology subgroups (0.17 logMAR in without vs 0.18 logMAR in with pathology; p=0.467). Intertest and test-retest accuracies were both around 0.00 logMAR (Supplementary Material, Figure S7). In our meta-analysis, differences in accuracy and precision between age groups and ocular health status were generally small. However, these analyses are limited to broad comparisons and limit the generalizability of our findings to specific age ranges, ocular diagnosis, or level of disease severity.

### Strength and Limitation

Several limitations should be considered. First, the imbalance in number of studies and effect estimates across testing strategies may have reduced the precision of subgroup comparisons and made sparely represented categories more sensitive to individual studies. Studies also contributed different number of effect estimates with varying sample size and sampling variance, resulting in unequal contribution to the pooled estimates. Although our multilevel models and cluster-robust inference accounted for dependence among effect estimates from the same study, uneven representation across testing strategies remains a limitation. This limitation was evident in the test-retest analysis. Several VA assessments, including Odysight(103), DigiVis(21), and myVisionTrack(110), have been developed for home self-testing or remote monitoring between clinic visits, but remote and unsupervised testing were each represented by only one study in our test-retest analysis. This limits our conclusions about repeatability under these intended conditions and highlight the need for further validation for longitudinal monitoring. Secondly, the methodological quality and clinical applicability of included studies was variable. Common patient-selection concerns included convenience or nonconsecutive recruitment, and many validation studies predominantly enrolled healthy young patients, limiting their applicability to boarder range of patient and ocular conditions encountered in clinical practice. Additionally, studies often did not clearly report whether examiners were masked to the results of the alternative test, which could influence test scoring, interpretation and potentially overestimate the agreement. These findings, together with substantial heterogeneity in intertest and test-retest estimates, highlight the need for rigorously designed validation studies using representative clinical population, prespecified testing protocols and masked assessment when feasible. Thirdly, we did not evaluate whether optotype design or the specific pairing of index and reference tests influence intertest agreement. Although ETDRS-based testing is widely regarded as the standardized reference for VA in clinical research(115), it was used as the comparator in only a little over half of the included intertest studies. Snellen, Landolt C, and Tumbling E charts were among other commonly used reference tests. The difference in optotype design, chart layout, crowding, scoring rules, and test-termination criteria may affect both systematic differences and the spread of differences between tests. Importantly, intertest LoA reflects the combined variability of the index and reference tests. Therefore, LoA half-width cannot be attributed solely to the performance of the index test. Consequently, heterogeneity in reference test selection may have contributed to the observed intertest variability in LoA half-width and potentially confounded comparisons across testing methodologies.

### Recommendations

We found that optotype sampling and response input method do not affect VA test precision but that testing site and level of supervision associated with great variability. The lack of difference in precision between fixed and adaptive sampling was likely due to use of fixed resolution of 0.10logMAR. This limits detection of finer resolution and constrained the smallest detectable change to 0.10logMAR. Likewise, a change would need to exceed 2 ETDRS lines to be considered a meaningful change beyond measurement noise. While ETDRS has been regarded as the gold-standard of VA measurement in the past decades, it may not be as sensitive to small changes in VA. While the addition of variability through different testing strategies, such as remote and unsupervised testings, add another layer of heterogeneity to VA assessments, contributing to the poor intertest agreement (0.19logMAR) and test-retest repeatability (0.14 logMAR). These findings suggest future development of VA assessment should aim to develop adaptive optotype sampling with higher test resolution under varying environment. Further research needs to determine whether these can improve test precision.

## Conclusion

This systematic review and meta-analysis comprehensively evaluated the accuracy and precision of VA assessments for both intertest and test-retest comparisons across four testing strategies, namely test site, optotype sampling, response input method, and level of supervision. Our findings suggest that precision is more susceptible than accuracy to sources of variability, especially in remote and unsupervised testing, but this result needs to be solidified with more supporting studies. With precision being consistent in between optotype sampling methods and response input methods reassure that these testing strategies might not be the main source of measurement variability. Instead, future investigation and development should aim at development of VA charts with finer resolution and finding their association with precision.

While adaptive tests showed no meaningful difference than fixed-scale tests, their ability to sample smaller step size without compromising precision suggest their potential in detecting meaningful changes beyond measurement noise. Hence, future VA development should utilize adaptive stimulus sampling and are recommended in clinical trials and for drug developments.

## Supplementary Information Captions

***Table S1. Reference visual acuity tests and the number of studies using each test.*** *ETDRS is the most used test, followed by Snellen and Landolt C*.

***Figure S1. Risk of bias and applicability concern analysis by QUADAS-2****. Each row represents an included study, and each column represents a QUADAS-2 domain. Risk of bias was assessed across patient-selection, index-test, reference standard, and flow-and-timing domains. Applicability concerns were assessed for the first three domains. The traffic lights represent high, low, or unclear concerns (red, green, and yellow, respectively)*.

***Figure S2 Summary of the QUADAS-2 risk of bias (RoB) and applicability concern analyses.*** *Bars show the proportional of included studies rated as having low, high, or unclear risk of bias or applicability concern within each domain. RoB was assessed across patient selection, index test, reference standard, and flow and timing. Applicability concerns were assessed for the first three domains. Low, high, and unclear RoB were represented in green, red, and yellow, respectively*.

***Figure S3. Overall Intertest Mean Bias Meta Analysis****. The forest plot represents individual effects contributed by each study. Left column reports first author and year of publication. Each square represents mean bias and 95%CI with corresponding numerical values on the right column. The dotted line represents no mean bias (0.00 logMAR). The diamond represents overall pooled systematic mean bias with 95%CI. The right column represents individual mean bias with 95%CI for each effect*.

***Figure S4. Overall Intertest LoA Half-Width Meta-Analysis****. The forest plot represents an individual effect contributed by each study. Left column reports first author and year of publication. Each square and horizontal bar represents LoA and 95%CI with corresponding numerical values on the right column. The diamond represents overall pooled LoA half-width with 95%CI*.

***Figure S5. Overall Test-retest Mean Bias Meta Analysis****. The forest plot represents an individual effect contributed by each study. Left column reports first author and year of publication. Each square represents test-retest mean bias and 95%CI with corresponding numerical values on the right column. The dotted line represents no bias (0.00 logMAR). The diamond represents overall pooled systematic mean bias with 95%CI*.

***Figure S6. Overall Test-retest CoR Meta Analysis****. The forest plot represents individual effect contributed by each study. Left column reports first author and year of publication. Each square represents CoR and 95%CI with corresponding numerical values on the right column. The diamond represents an overall CoR with 95%CI*.

***Table S2. Intertest mean bias by testing strategies****. Mean bias differed between adaptive and fixed test resolutions, but the between-group comparison did not reach statistical significance (p=0.060). None of the four testing strategies displayed a statistically significant difference in mean bias between its paired-wise comparisons (all p>0.05). The numbers of effects were unbalanced for clinic vs remote testing (k=106 vs k=9) and supervised vs unsupervised (k=107 vs k=7). A residual τ_total_ of 0.07 logMAR indicates that underlying mean bias varied around their subgroup estimates with a standard deviation of approximately 0.07 logMAR (3.5 ETDRS letters). Estimates are expressed in logMAR. Separate moderator models were fitted for each testing strategy*.

***Table S3. Intertest LoA half-width by testing strategies****. LoA half-width differed significantly between clinic and remote testing (p=0.030), but the number of contributing effects was unbalanced (k=99 vs k=8). No significant difference was detected for the other testing strategies. Residual heterogeneity was reported as τ_total_ on the natural-log scale and as e τ_total_ a multiplicative factor on a ratio scale. Both describe variation remaining among study effects after accounting for the differences between groups defined by each testing strategies. Pooled estimates were presented in logMAR values. Separate moderator models were fitted for each testing strategies*.

***Table S4. Test-retest mean bias by testing strategies****. Test-retest mean bias differed between examiner and self-input VA assessments (p=0.04). No statistical significant difference was observed between fixed and adaptive test resolutions. Residual of 0.02 logMAR indicates that underlying test-retest mean bias varied around their subgroup estimates with a standard deviation of approximately 0.02 logMAR (1 ETDRS letters). Estimates are expressed in logMAR values. Separate moderator models were fitted for each testing strategy. Test site and supervision were not evaluated as moderators because the remote and unsupervised categories each contained only one study*.

***Table S5. Test-retest CoR by testing strategies****. CoR differed significantly between examiner and self-input VA assessments (p=0.030). No significant difference was detected between fixed and adaptive test resolutions (p=0.098). Residual heterogeneity was reported as τ_total_ on the natural-log scale and as e τ_total_*, *a multiplicative factor on a ratio scale. Both describe variation remaining among study effects after accounting for the differences between groups defined by each testing strategy. Pooled estimates were presented in logMAR values. Separate moderator models were fitted for each testing strategy. Test site and supervision were not evaluated as moderators because the remote and unsupervised categories each contained only one study*.

***Figure S7. Intertest (top) and test-retest (bottom) accuracy and precision outcomes by age subgroups****. A statistically significant difference was detected between intertest accuracy between adult and children (p=0.008). No other statistically significant difference was observed in other accuracy and precision measures. Dotted line in mean bias plot represents no systematic mean bias (0.00 logMAR)*.

***Figure S8. Intertest (top) and test-retest (bottom) accuracy and precision outcomes by pathology subgroups****. The LoA half-width and CoR for each subgroup was consistent with the overall pooled estimates. No statistically significant difference was observed in accuracy and precision measures. Dotted line in mean bias plot represents no systematic mean bias (0.00 logMAR)*.

## Registration and Protocol

Review was not registered and protocol was not prepared.

## Acknowledgements

Portions of the study were presented as a poster at ARVO 2025.

## Author Contributions

AB and JS conceptualized the study. CC and JS developed the methodology, designed and executed the literature search. CC performed study screening, data curation. JS and CC performed the review and risk-of-bias assessment of selected studies. CC conducted the formal analysis and prepared the visualization outputs. CC drafted the original manuscript. JS and AB contributed to writing—review and editing. AB and JS provided supervision and project administration. All authors reviewed and approved the final manuscript.

## AI Statement

ChatGPT-4 (OpenAI) was used to assist with grammar and language refinement, preparation of analysis software, generation of figures 1-2, as well as to help refine preliminary search terms during development of the literature search strategy. All AI-assisted outputs were reviewed, verified, and edited by the authors, who take full responsibility for the final manuscript.

## Data Availability

Data will be available on DRYAD at DOI https://doi.org/10.5061/dryad.3ffbg7b24.

