## Supplementary Materials for "Accuracy and precision of visual acuity tests: A systematic review and meta-analysis across testing strategies"

**Full Research Syntax**

**Inclusion/Exclusion Criteria**

**Table S1. Reference visual acuity tests and the number of studies using each test**

**Table S2. Intertest mean bias by testing strategies**

**Table S3. Intertest LoA half-width by testing strategies**

**Table S4. Test-retest mean bias by testing strategies**

**Table S5. Test-retest CoR by testing strategies.**

**Full Search Syntax**

PubMed:

("visual acuity" OR "resolution" OR "vision acuity") AND (snellen OR landolt OR ETDRS OR Tumbling) AND (conventional OR lightbox OR printed OR automated OR digital OR computerized OR autonomous OR unsupervised OR self-assessed OR self-administered OR self-measured OR digital OR mobile app OR application OR smartphone OR remote OR letter chart OR chart OR eye chart) AND (tools OR assessment OR protocol OR test OR method) AND (validation OR reliability OR repeatability OR bland altman OR comparison)

Google Scholar:

validation or comparison or repeatability or reliability or comparison "visual acuity" or "eye chart"

Embase:

('visual acuity' OR 'resolution' OR 'vision acuity') AND ('snellen' OR 'landolt' OR 'etdrs' OR 'tumbling') AND ('conventional' OR 'lightbox' OR 'printed' OR 'automated' OR 'computerized' OR 'autonomous' OR 'unsupervised' OR 'self-assessed' OR 'self-administered' OR 'self-measured' OR 'digital' OR 'mobile app' OR 'application' OR 'smartphone' OR 'remote' OR 'letter chart' OR 'chart' OR 'eye chart') AND ('tools' OR 'assessment' OR 'protocol' OR 'test' OR 'method') AND ('validation' OR 'reliability' OR 'repeatability' OR 'bland altman' OR 'comparison')

Scopus:

('visual acuity' OR 'resolution' OR 'vision acuity') AND ('snellen' OR 'landolt' OR 'etdrs' OR 'tumbling') AND ('conventional' OR 'lightbox' OR 'printed' OR 'automated' OR 'computerized' OR 'autonomous' OR 'unsupervised' OR 'self-assessed' OR 'self-administered' OR 'self-measured' OR 'digital' OR 'mobile app' OR 'application' OR 'smartphone' OR 'remote' OR 'letter chart' OR 'chart' OR 'eye chart') AND ('tools' OR 'assessment' OR 'protocol' OR 'test' OR 'method') AND ('validation' OR 'reliability' OR 'repeatability' OR 'bland altman' OR 'comparison')

**Inclusion/Exclusion Criteria**

Include studies that:

- Enrolled human participants undergoing subjective visual acuity testing, regardless of age or ocular health
- Evaluate optotype-based test requiring a behavioral response from the participants
- Reported either
  - an intertest comparison between an index VA test and a reference VA test, or
  - a test-retest comparison involved repeated measurements of the same VA test
- Used Bland-Altman analysis or reported sufficient data to determine
  - Mean bias and 95% limits of agreement for intertest comparison, or
  - Mean bias and coefficient of repeatability of test-retest comparison
- Used an observational or interventional study design that provided eligible agreement data, including cross-sectional, cohort, and diagnostic-comparison.
- Articles published in English
- Articles published between 1 January 1983 to 02 December 2024

Exclude studies that:

- Evaluated objective or physiological estimates of VA without a subjective participant response, such as visual evoked potential or eye-tracking-measured acuity
- Evaluated non-optotype based VA
- Compared VA before and after intervention (treatment, surgery, induced blur)
- Reported only correlation coefficient, sensitivity, specificity, mean VA values or other metrics without Bland-Altman agreement data.
- Published as case reports, reviews, editorials, letters without original data, study protocols, conference abstracts, or abstracts without accessible full-text

| **Reference test used** | **Studies, n** |
| --- | --- |
| ETDRS chart | 44 |
| Snellen chart | 19 |
| Landolt C chart | 3 |
| LEA Symbols test | 3 |
| Rosenbaum near card | 3 |
| Kay Pictures test | 1 |
| Baile-Lovie acuity chart | 1 |
| TNO Landolt C acuity chart | 1 |
| COMPlog assessment | 1 |

***Table S1****.* ***Reference visual acuity tests and the number of studies using each test.*** *ETDRS is the most used test, followed by Snellen and Landolt C.*

| Intertest Mean Bias by Testing Strategies | | | | | | | |
| --- | --- | --- | --- | --- | --- | --- | --- |
| Testing Strategies | **Level** | **Studies, n** | **Effects, k** | **Pooled estimate (95%CI)** | **Between-group Difference (95%CI)** | **HTZ Moderator test, F (df1, df2); P** | **Residual τ** |
| Test Site | Clinic | 64 | 106 | 0.01  (-0.01, 0.03) | Remote - Clinic  0.00 (-0.08, 0.08) | *F* (1, 5.42) = 0.01, p=0.941 | 0.07 |
|  | Remote | 6 | 8 | 0.01  (-0.07, 0.09) |  |  |  |
| Optotype Sampling | Fixed | 37 | 53 | 0.02  (0.00, 0.05) | Adaptive - Fixed  -0.02 (-0.04, 0.00) | *F* (1, 32.89) = 3.79, p=0.060 | 0.07 |
|  | Adaptive | 38 | 61 | 0.00  (-0.01, 0.02) |  |  |  |
| Response Input | Examiner | 51 | 85 | 0.01  (-0.01, 0.03) | Self - Examiner  -0.01 (-0.04, 0.02) | *F* (1, 13.63) = 0.50, p=0.493 | 0.07 |
|  | Self | 21 | 29 | 0.00  (-0.02, 0.03) |  |  |  |
| Supervision | Supervised | 64 | 107 | 0.01  (0.00, 0.03) | Unsupervised - Supervised  -0.02 (-0.11, 0.08) | *F* (1, 4.53) = 0.21, p=0.666 | 0.07 |
|  | Unsupervised | 5 | 7 | -0.01  (-0.10, 0.09) |  |  |  |

***Table S2. Intertest mean bias by testing strategies.*** *Mean bias differed between adaptive and fixed sampling, but the between-group comparison did not reach statistical significance (p=0.060). None of the four testing strategies displayed a statistically significant difference in mean bias between its paired-wise comparisons (all p>0.05). The numbers of effects were unbalanced for clinic vs remote testing (k=106 vs k=9) and supervised vs unsupervised (k=107 vs k=7). A residual 𝜏_total_ of 0.07 logMAR indicates that underlying mean bias varied around their subgroup estimates with a standard deviation of approximately 0.07 logMAR (3.5 ETDRS letters). Estimates are expressed in logMAR. Separate moderator models were fitted for each testing strategy.*

| Intertest LoA Half-Width by Testing Strategies | | | | | | | | |
| --- | --- | --- | --- | --- | --- | --- | --- | --- |
| Testing Strategies | **Level** | **Studies, n** | **Effects, k** | **Pooled estimate (95%CI)** | **Between-group Ratio (95%CI)** | **HTZ Moderator test, F (df1, df2); P** | **Residual Heterogeneity**  **(τ_total_)** | **Multiplicative factor**  (e^τtotal^) |
| Test Site | Clinic | 61 | 99 | 0.19  (0.17, 0.21) | Remote/Clinic  1.46 (1.05, 2.02) | *F* (1, 5.56) = 8.42, p=0.030 | 0.49 | 1.63 |
|  | Remote | 6 | 8 | 0.27  (0.20, 0.38) |  |  |  |  |
| Optotype  Sampling | Fixed | 37 | 52 | 0.18  (0.16, 0.21) | Adaptive/Fixed  1.10 (0.93, 1.32) | *F* (1, 39.01) = 1.28, p=0.266 | 0.50 | 1.65 |
|  | Adaptive | 35 | 55 | 0.20  (0.18, 0.24) |  |  |  |  |
| Response Input | Examiner | 48 | 78 | 0.19  (0.17, 0.22) | Self/Examiner  0.99 (0.77, 1.28) | *F* (1, 16.60) = 0.01, p=0.925 | 0.51 | 1.66 |
|  | Self | 21 | 29 | 0.19  (0.15, 0.25) |  |  |  |  |
| Supervision | Supervised | 61 | 100 | 0.19  (0.17, 0.21) | Unsupervised/  emSupervised  1.37 (0.90, 2.07) | *F* (1, 4.46) = 4.04, p=0.108 | 0.50 | 1.64 |
|  | Unsupervised | 5 | 7 | 0.26  (0.17, 0.39) |  |  |  |  |

***Table S3. Intertest LoA half-width by testing strategies****. LoA half-width differed significantly between clinic and remote testing (p=0.030), but the number of contributing effects was unbalanced (k=99 vs k=8). No significant difference was detected for the other testing strategies. Residual heterogeneity was reported as 𝜏_total_ on the natural-log scale and as e ^𝜏^_total,_ a multiplicative factor on a ratio scale. Both describe variation remaining among study effects after accounting for the differences between groups defined by each testing strategies. Pooled estimates were presented in logMAR values. Separate moderator models were fitted for each testing strategies.*

| Test-retest Mean Bias by Testing Strategies | | | | | | | |
| --- | --- | --- | --- | --- | --- | --- | --- |
| Testing strategy | **Level** | **Studies, n** | **Effects, k** | **Pooled estimate (95%CI)** | **Between-group Difference (95%CI)** | **HTZ Moderator test, F (df1, df2); P** | **Residual τ** |
| Test Site | Clinic | 40 | 89 | 0.01  (0.00, 0.01) | - | *-* | - |
|  | Remote | - | - | - | - | *-* | - |
| Optotype Sampling | Fixed | 23 | 41 | 0.01  (0.00, 0.01) | Adaptive - Fixed  0.00 (-0.01, 0.01) | *F* (1,24.41) = 0.01, p=0.091 | 0.02 |
|  | Adaptive | 28 | 49 | 0.01  (0.00, 0.01) |  |  |  |
| Response Input | Examiner | 32 | 69 | 0.01  (0.00, 0.01) | Self - Examiner  -0.01 (-0.02, 0.00) | *F* (1,13.30) = 5.24, p=0.039 | 0.02 |
|  | Self | 15 | 21 | 0.00  (-0.01 0.01) |  |  |  |
| Supervision | Supervised | 40 | 89 | 0.01  (0.00, 0.01) | - | *-* | - |
|  | Unsupervised | - | - | - | - | *-* | - |

***Table S4. Test-retest mean bias by testing strategies.*** *Test-retest mean bias differed between examiner and self-input VA assessments (p=0.04). No statistical significant difference was observed between fixed and adaptive sampling. Residual 𝜏 of 0.02 logMAR indicates that underlying test-retest mean bias varied around their subgroup estimates with a standard deviation of approximately 0.02 logMAR (1 ETDRS letters). Estimates are expressed in logMAR values. Separate moderator models were fitted for each testing strategy. Test site and supervision were not evaluated as moderators because the remote and unsupervised categories each contained only one study.*

| Test-retest CoR by Testing Strategies | | | | | | | | |
| --- | --- | --- | --- | --- | --- | --- | --- | --- |
| Test Strategies | **Level** | **Studies, n** | **Effects, k** | **Pooled estimate (95%CI)** | **Between-group Ratio (95%CI)** | **HTZ Moderator test, F (df1, df2); P** | **τ_total_** | **Multiplicative factor**  (e^τtotal^) |
| Test site | Clinic | 44 | 100 | 0.14  (0.12, 0.16) | - | *-* | - | - |
|  | Remote | - | - | - |  |  |  |  |
| Optotype Sampling | Fixed | 27 | 50 | 0.13  (0.11, 0.16) | Adaptive/Fixed  1.20 (0.96, 1.50) | *F* (1,28.67) = 2.91, p=0.098 | 0.53 | 1.70 |
|  | Adaptive | 29 | 51 | 0.15  (0.13, 0.18) |  |  |  |  |
| Response Input | Examiner | 36 | 80 | 0.13  (0.11, 0.15) | Self/Examiner  1.39 (1.04, 1.85) | *F* (1,13.83) = 5.83, p=0.030 | 0.53 | 1.69 |
|  | Self | 15 | 21 | 0.18  (0.14, 0.23) |  |  |  |  |
| Supervision | Supervised | 44 | 100 | 0.14  (0.12, 0.16) | - | - | - | - |
|  | Unsupervised | - | - | - |  |  |  |  |

***Table S5. Test-retest CoR by testing strategies****. CoR differed significantly between examiner and self-input VA assessments (p=0.030). No significant difference was detected between fixed and adaptive sampling (p=0.098). Residual heterogeneity was reported as 𝜏_total_ on the natural-log scale and as e ^𝜏^_total,_ a multiplicative factor on a ratio scale. Both describe variation remaining among study effects after accounting for the differences between groups defined by each testing strategy. Pooled estimates were presented in logMAR values. Separate moderator models were fitted for each testing strategy. Test site and supervision were not evaluated as moderators because the remote and unsupervised categories each contained only one study.*
