## Supplementary figures and images for "Accuracy and precision of visual acuity tests: A systematic review and meta-analysis across testing strategies"

### Supplement Material, Figure S7

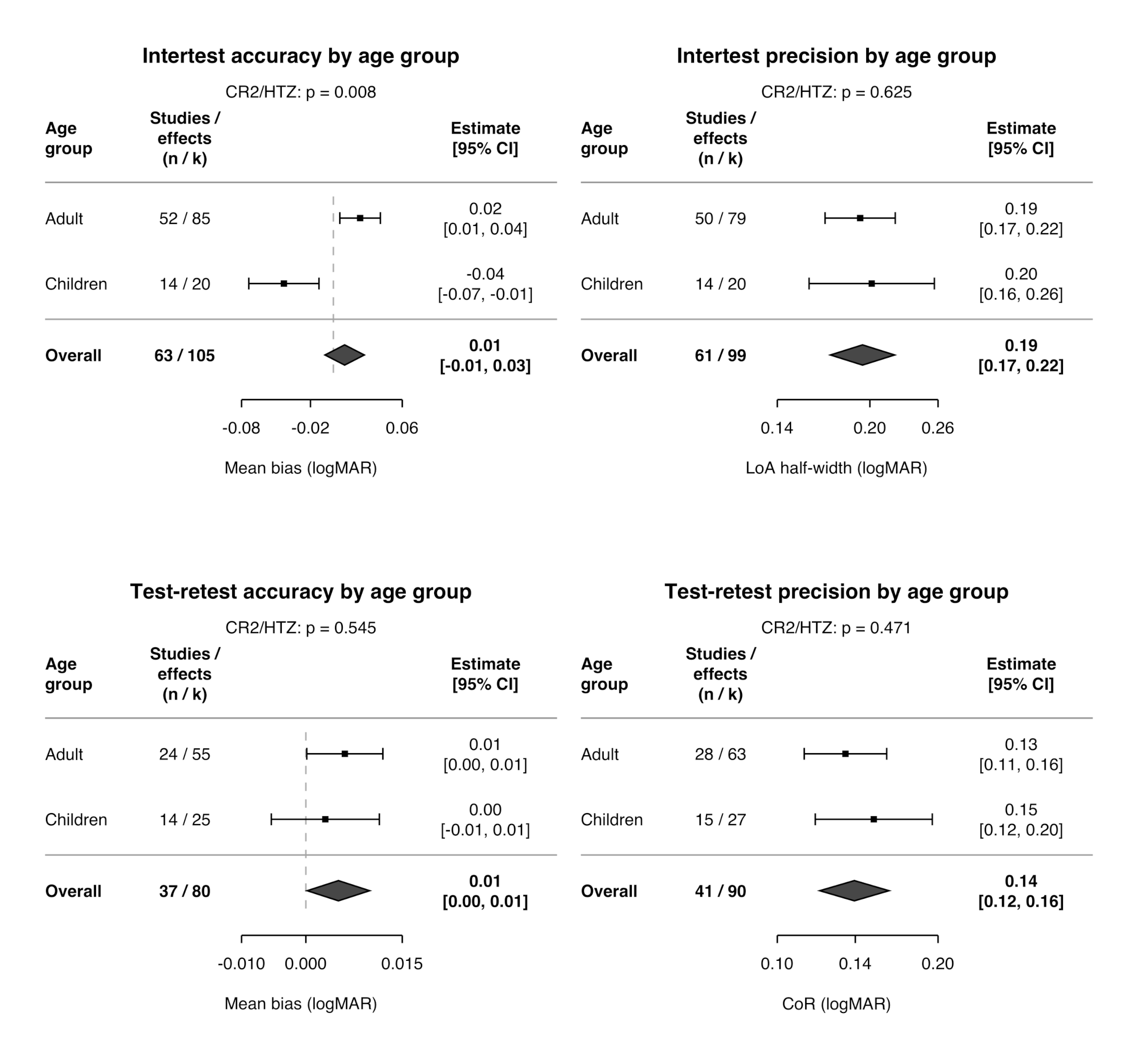

### Supplement Material, Figure S8

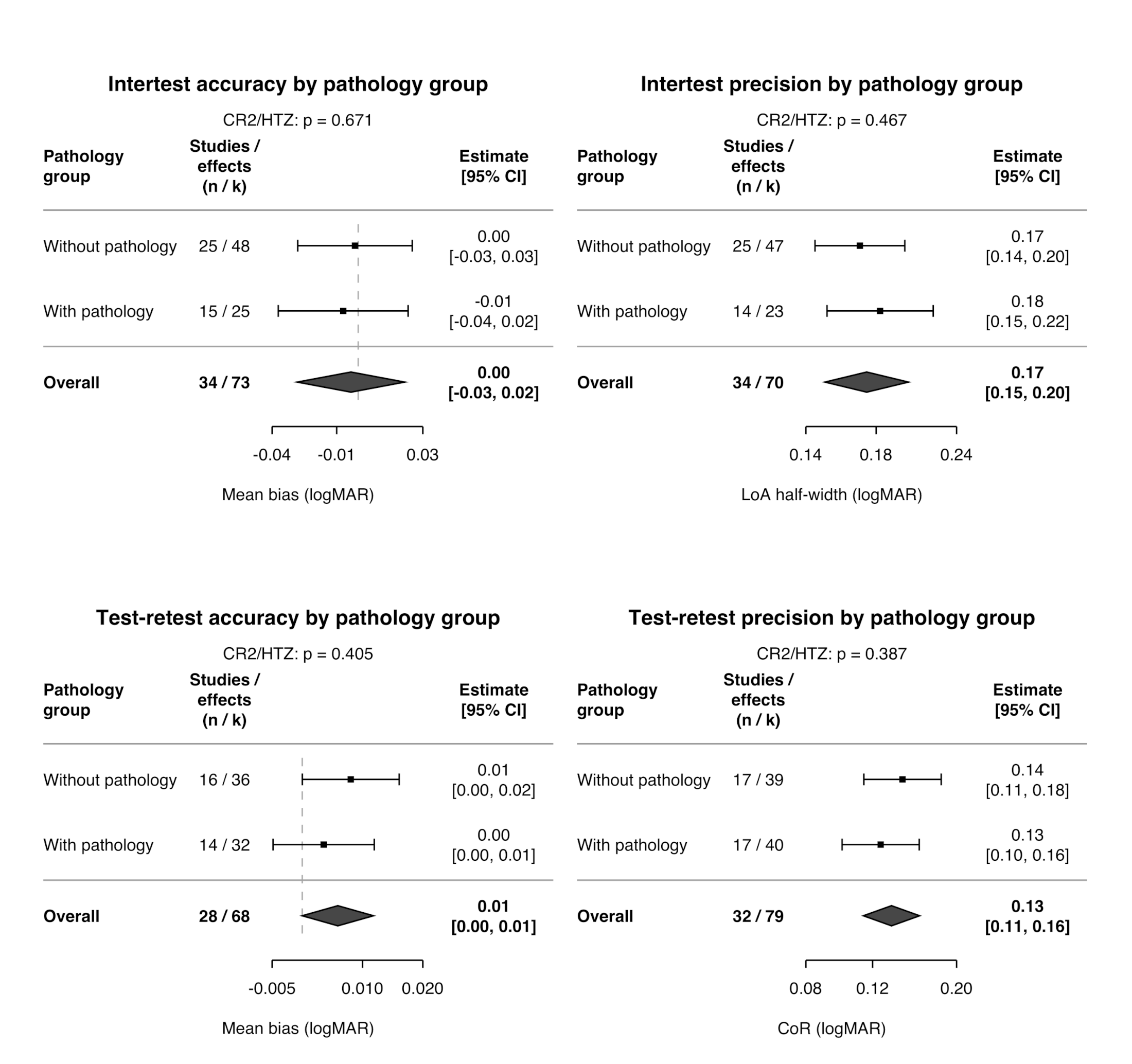

### Supplement Materials, Figure S1

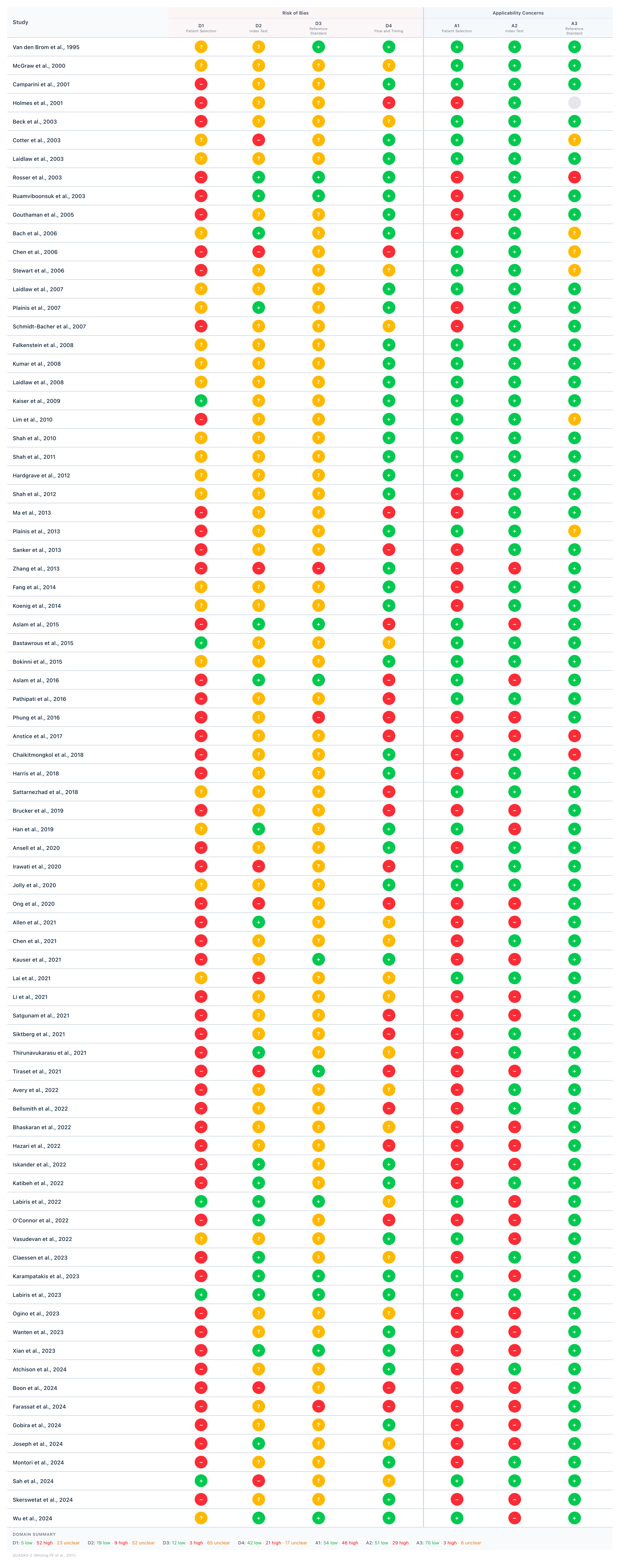

### Supplement Materials, Figure S2

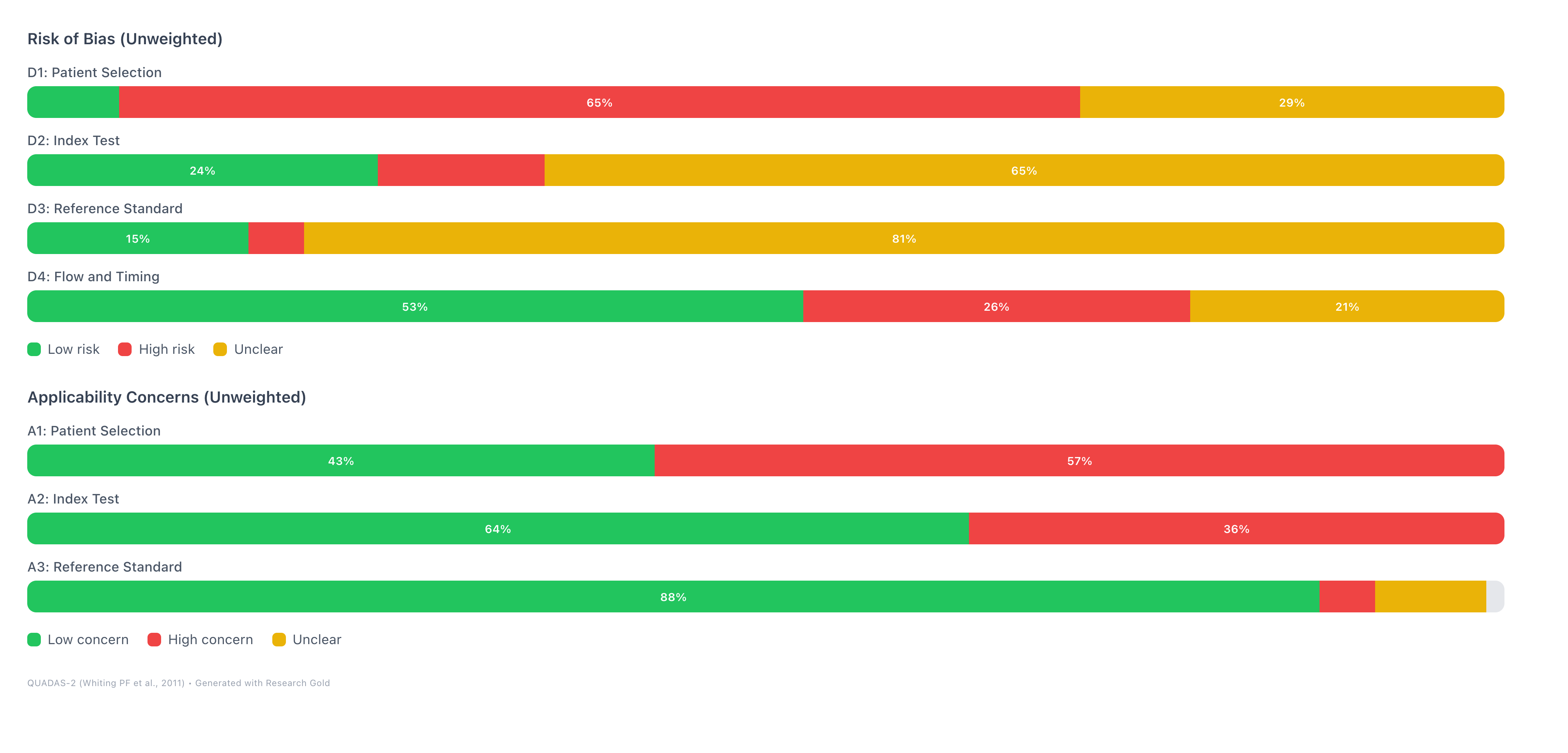

### Supplementary Material Figure S3

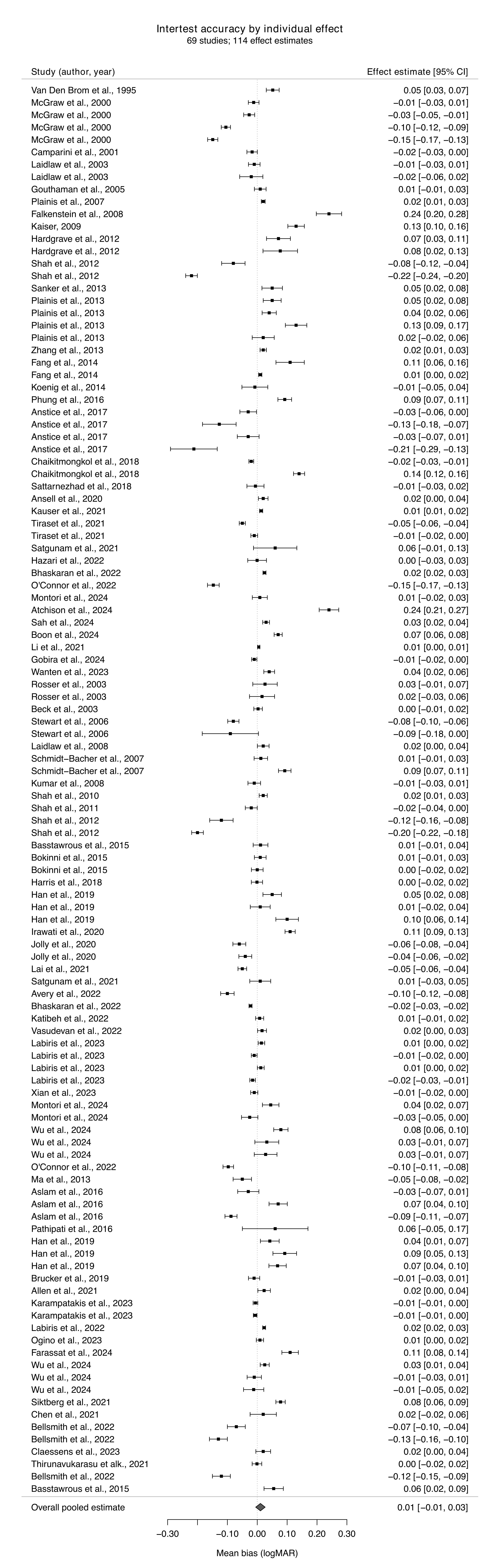

### Supplementary Material Figure S4

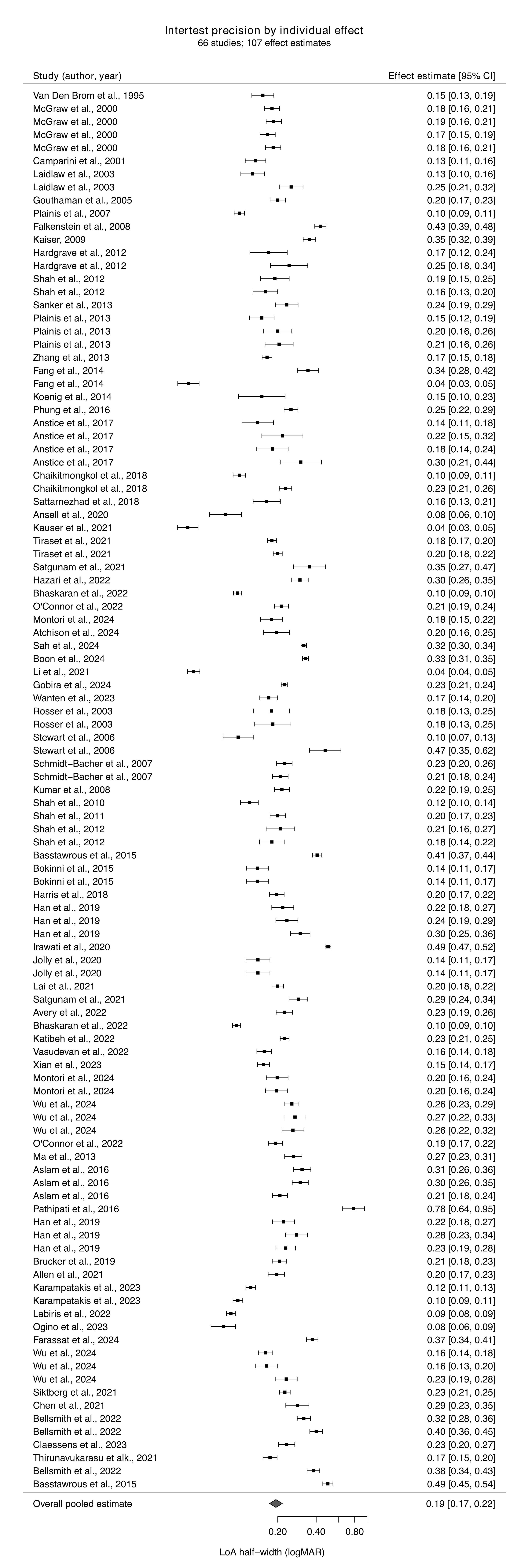

### Supplementary Material Figure S5

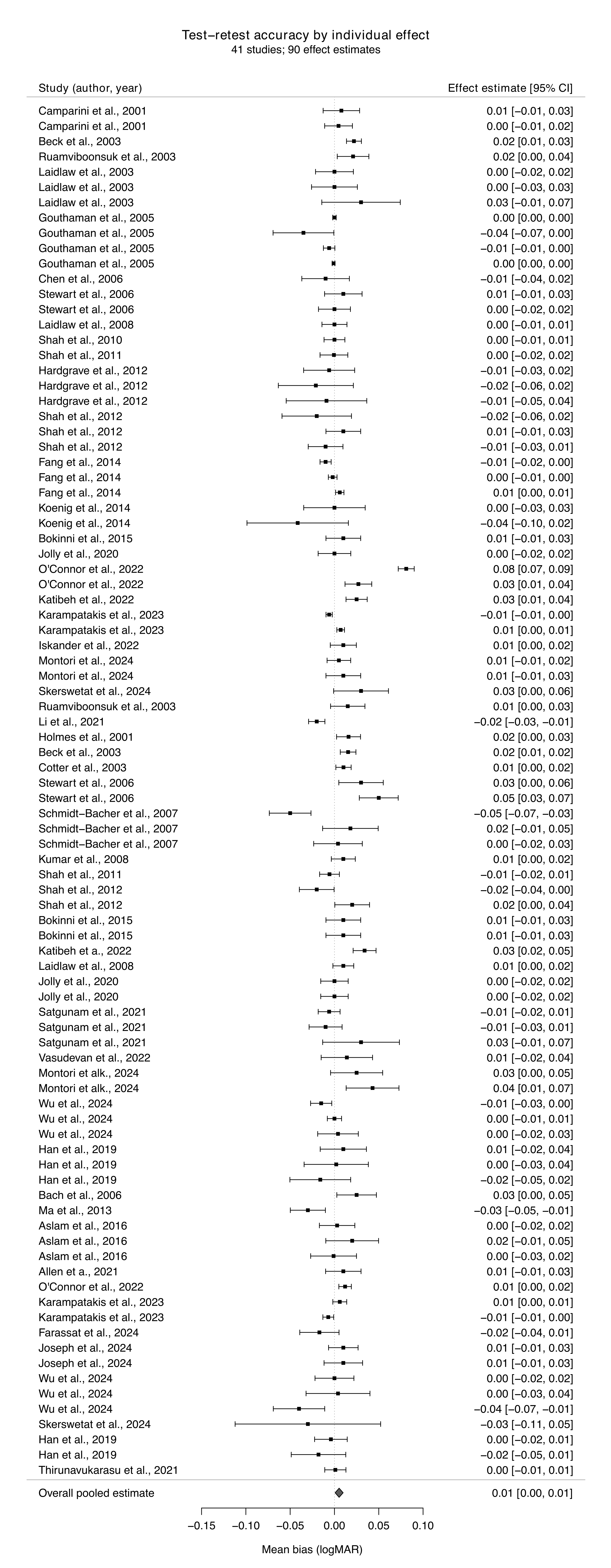

### Supplementary Material Figure S6

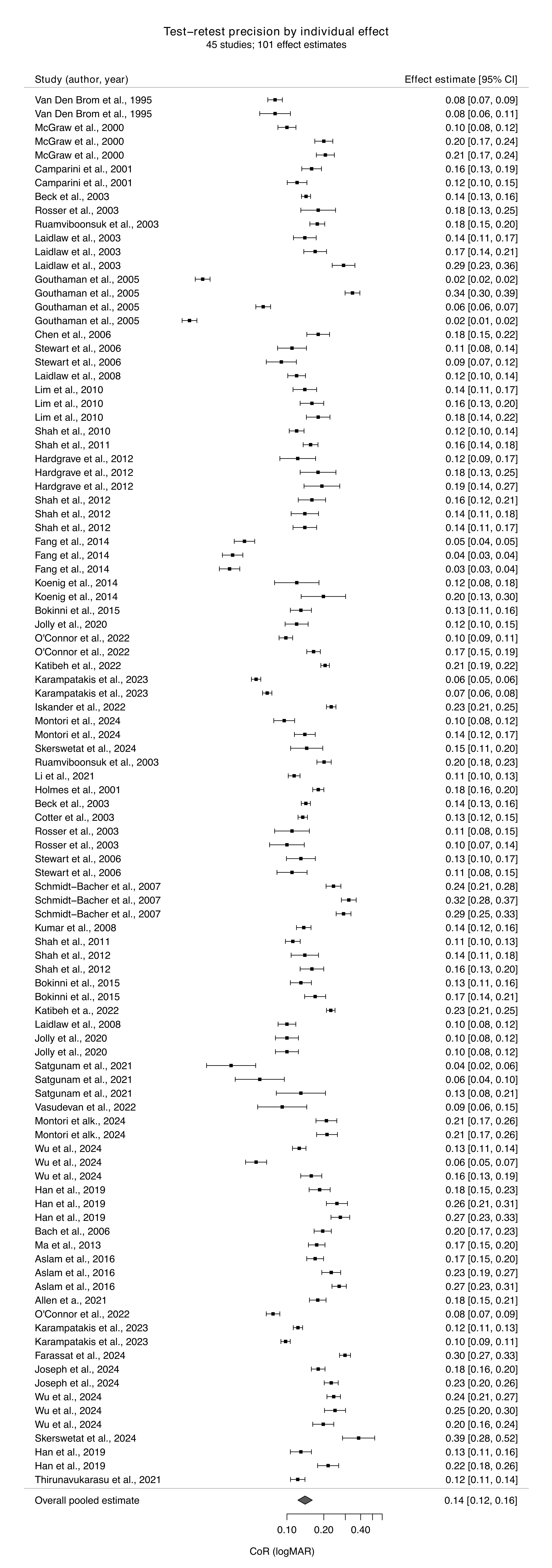
